# Analysis of daily reproduction rates of COVID-19 using Current Health Expenditure as Gross Domestic Product percentage (CHE/GDP) across countries

**DOI:** 10.1101/2021.08.27.21262737

**Authors:** Kayode Oshinubi, Mustapha Rachdi, Jacques Demongeot

## Abstract

**Background:** Impact and severity of coronavirus pandemic on health infrastructure vary across countries. We examine the role percentage health expenditure plays in various countries in terms of their preparedness and see how countries improved their public health policy in the first and second wave of the coronavirus pandemic;

**Methods:** We considered the infectious period during the first and second wave of 195 countries with their Current Health Expenditure as Gross Domestic Product percentage (CHE/GDP). Exponential model was used to calculate the slope of the regression line while the ARIMA model was used to calculate the initial autocorrelation slope and also to forecast new cases for both waves. The relationship between epidemiologic and CHE/GDP data was used for processing ordinary least square multivariate modeling and classifying countries into different groups using PC analysis, K-means and Hierarchical clustering;

**Results:** Results show that some countries with high CHE/GDP improved their public health strategy against virus during the second wave of the pandemic;

**Conclusions:** Results revealed that countries who spend more on health infrastructure improved in the tackling of the pandemic in the second wave as they were worst hit in the first wave. This research will help countries to decide on how to increase their CHE/GDP in order to tackle properly other pandemic waves of the present Covid-19 outbreak and future diseases that may occur. We are also opening up a debate on the crucial role socio-economic determinants play during the exponential phase of the pandemic modelling.

## 1. Introduction

So far, most countries have experienced at least two peaks of the Covid-19 pandemic and it is necessary to look at both waves and then derive the best conclusion on the efficacy of outlook during these both waves. Health officials, scientists and those involved in the modelling of the pandemic have made a lot of suggestions from the day the first case has been recorded in Wuhan, China. The Current Health Expenditure as Gross Domestic Product percentage (CHE/GDP) is key to different countries’ preparedness to respond for curtailing the pandemic even though it is general belief that no one was prepared during the first wave of the pandemic as most developed nations were worst hit and the death toll increased exponentially.

Our goal is to correlate the maximum basic reproduction number R_0_ of both waves with the CHE/GDP. In order to holistically approach this subject, we used many diverse regression tools and also developed some clustering strategies across all countries considered. The results are key in order to protect lives and improve health infrastructure in the future even though we know that the pandemic is still evolving in different countries.

## 2. Materials and Methods

### 2.1. Materials

#### 2.1.1. Variables

The variables used for this research are seven in total. The maximum basic reproduction number R_0_ for first and second waves is chosen during the exponential phase of all countries considered. The exponential and autocorrelation slopes are calculated using 100 days from the start of a wave depending on the date a particular country recorded their first case between February and August 2020 while also 100 days was used to calculate for the second wave between October 15 2020 to January 22 2021 for all countries considered. The opposite of initial autocorrelation slope was averaged on six days. CHE/GDP was collated from World Bank data [1]. The deterministic R_0_ was drafted from previous research [2] and it was calculated as the Malthusian growth parameter during the exponential phase of both waves across countries. The daily new cases were drafted from Worldometers^®^ [3] and Renkulab^®^ [4] databases.

### 2.2. Methods

#### 2.2.1. Exponential and ARIMA model

The exponential model is given as y = ae^bx^, where y is the daily number of new cases, x is the number of days, b is the slope and a is a constant, and the log format can be written as log y = log a + bx.

ARIMA modelling has been introduced by N. Wiener for prediction and forecasting [5]. Its parametric approach assumes that the underlying stationary stochastic process of the COVID-19 new daily cases N(t) can be described by a small number of parameters using the autoregressive ARIMA model N(t) = Σ_i=1,s_ a(i) N(i) + W(t), where W is a random residue whose variance is to minimize. The autocorrelation analysis is done by calculating the correlation A(k) between the N(t)’s and the N(t − k)’s (t belonging to a moving time window) by using the formula

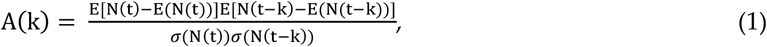

where E denotes the expectation and σ the standard deviation. The autocorrelation function A allows examining the serial dependence of the N(t)’s. We used ARIMA form of (6, 1, 0), which we think is the best for the modelling.

#### 2.2.2. Clustering methodology

Clustering is a branch of machine learning which is called ‘unsupervised learning’ and is frequently utilized to classify biomedical data. We used three classical clustering methods, K-means, PCA (Principal Component Analysis) and Hierarchical clustering [6]. K-means clustering chooses a priori the number of clusters and starts out with random centroids while Hierarchical clustering starts with every point in dataset as a cluster, then finds the two closest points and combines them into clusters, the process being repeated until appears a big giant cluster and it then creates a dendrogram.

Principal component analysis (PCA) also helps to cluster data points and it is also one of dimension reduction techniques because each variable has a different dimension. It allows us to summarize and visualize the information in a data set described by multiple inter-correlated variables. PCA is used to extract the important information from variables in the dataset and to express this information as a set of few new variables called principal components (PC’s).

#### 2.2.3. Linear and Polynomial Regression

Linear regression models use some historic data (100 days infectivity period in our case) of independent and dependent variables (CHE/GDP) and consider a linear relationship between both while polynomial regression models use a similar approach but the dependent variable is modeled as a degree n (6≥n≥2) polynomial in x.

#### 2.2.4. Multivariate Ordinary least square method

Multivariate least square method allows us to test much more complex relations between variables. It can be can be represented as follows:

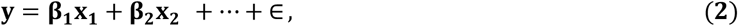

where **β**_**1**_, **β**_**2**_, … are coefficients or weights, ∈ is the residual noise, y is the dependent variable and **x**_**1**_, **x**_**2**_, … are the independent variables.

## 3. Results

### 3.1. Autocorrelation Slope

#### 3.1.1. Parabolic and cubic regression

**Figure 1.**
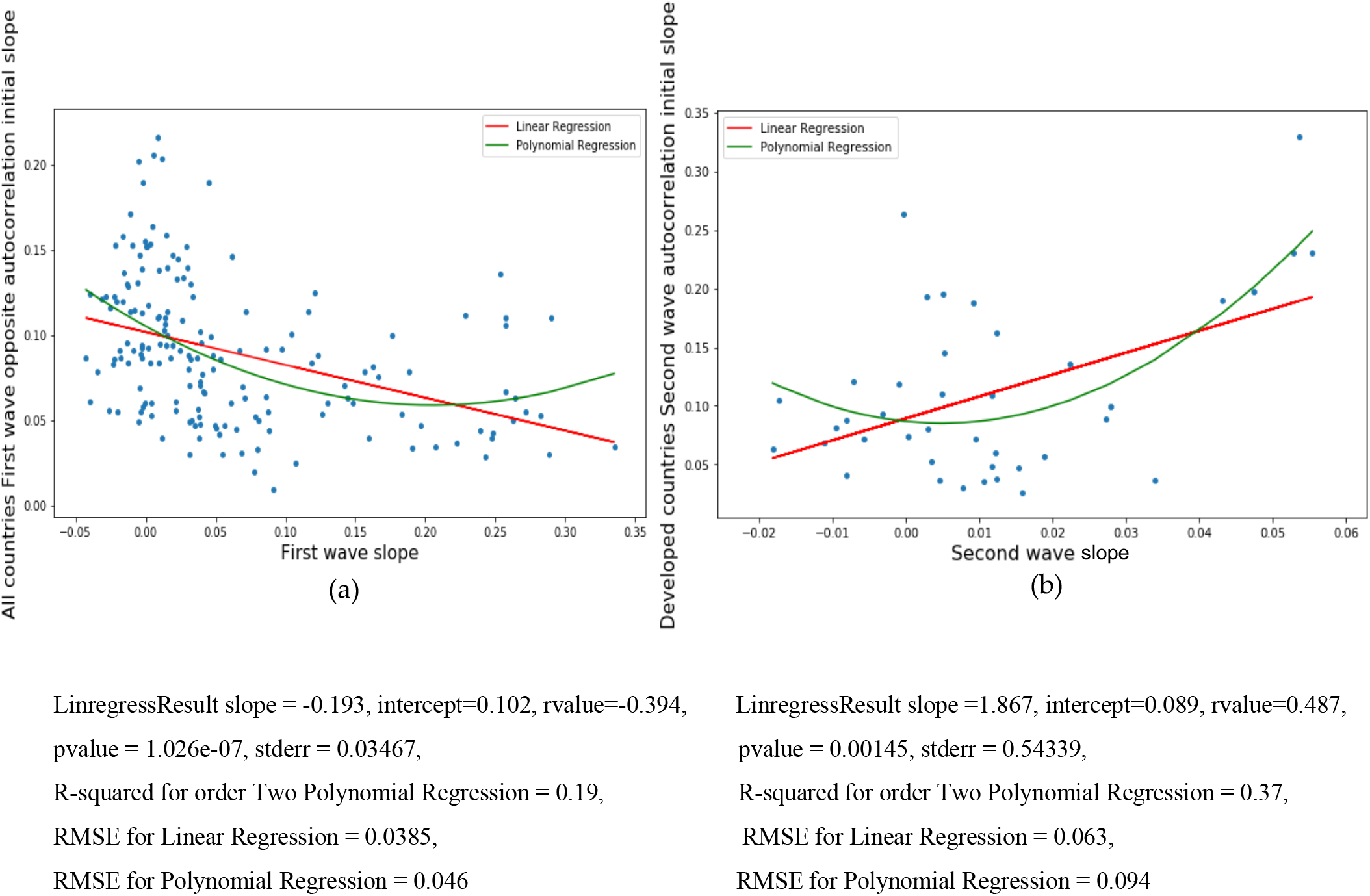
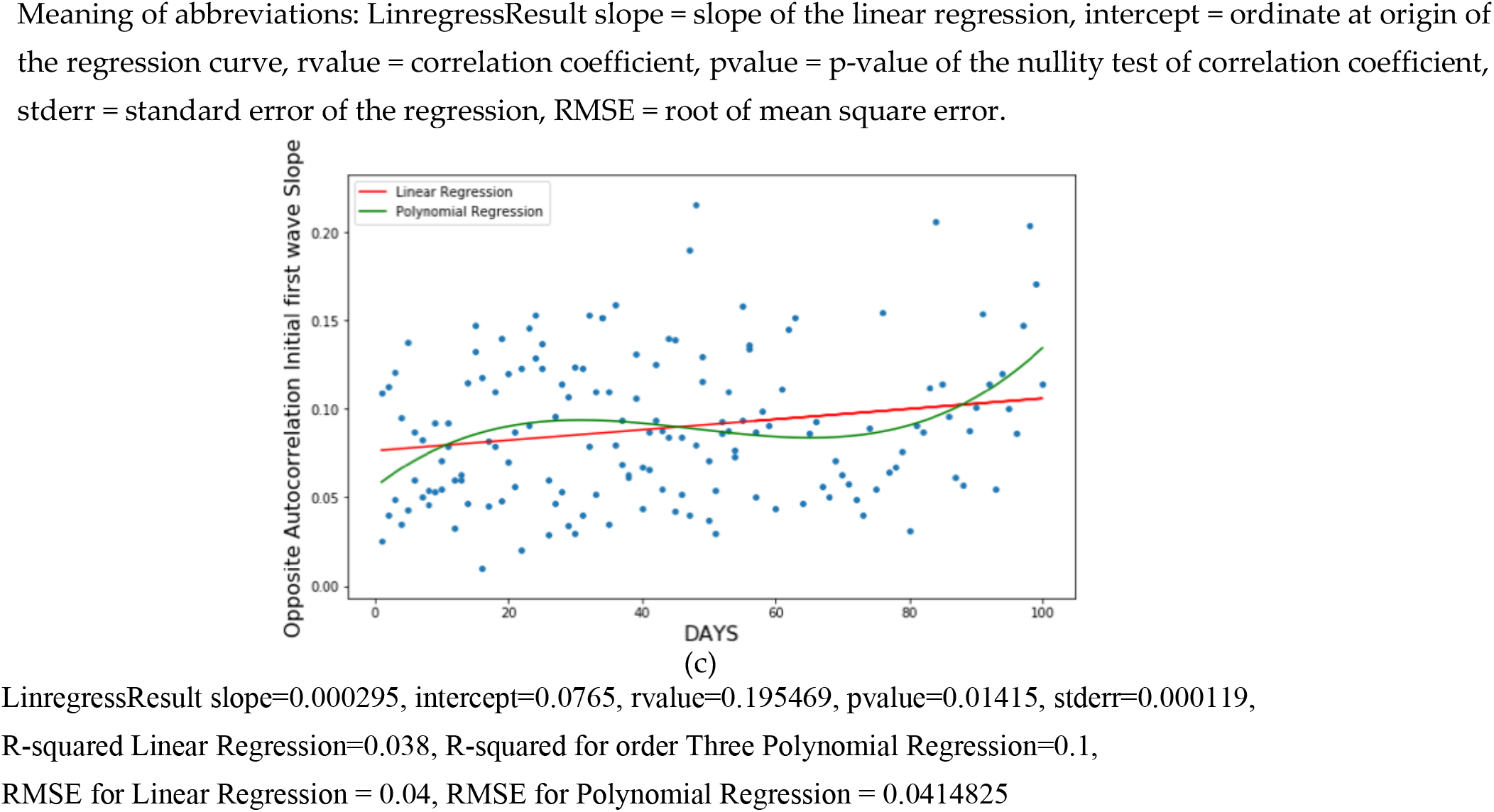
Linear (in red) and parabolic or cubic (in green) regression plots of the opposite of the initial autocorrelation slope vs (a) first wave exponential regression slope for all countries, (b) second wave exponential regression slope for developed countries and (c) days from the start of the first wave observed in China for all countries.

#### 3.1.2. Quartic regression

**Figure 2.**
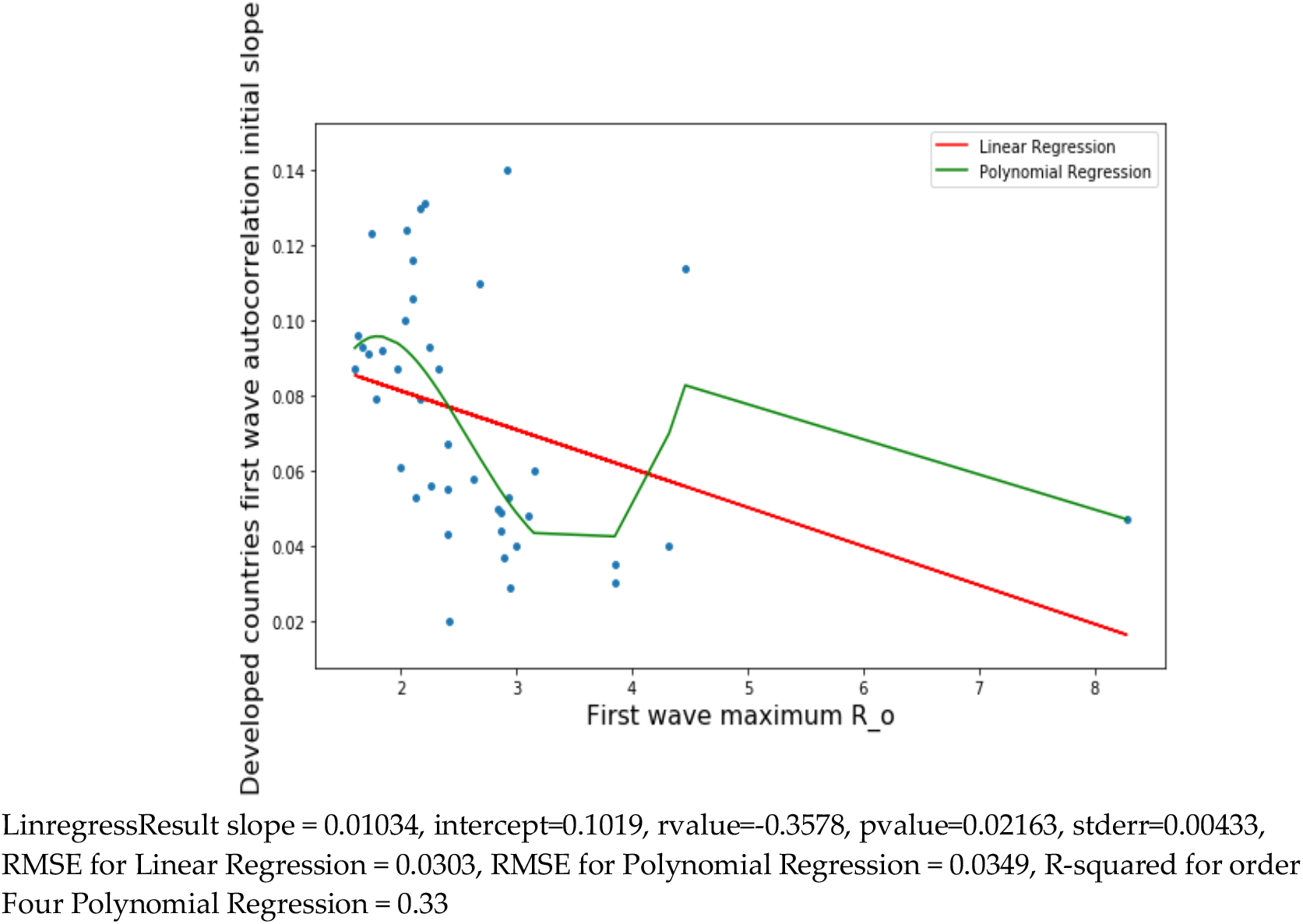
Linear (in red) and quartic (in green) regression plots of the opposite of the initial autocorrelation slope of the first wave vs first wave maximum R_0_ for developed countries.

#### 3.1.3. Sextic regression

**Figure 3.**
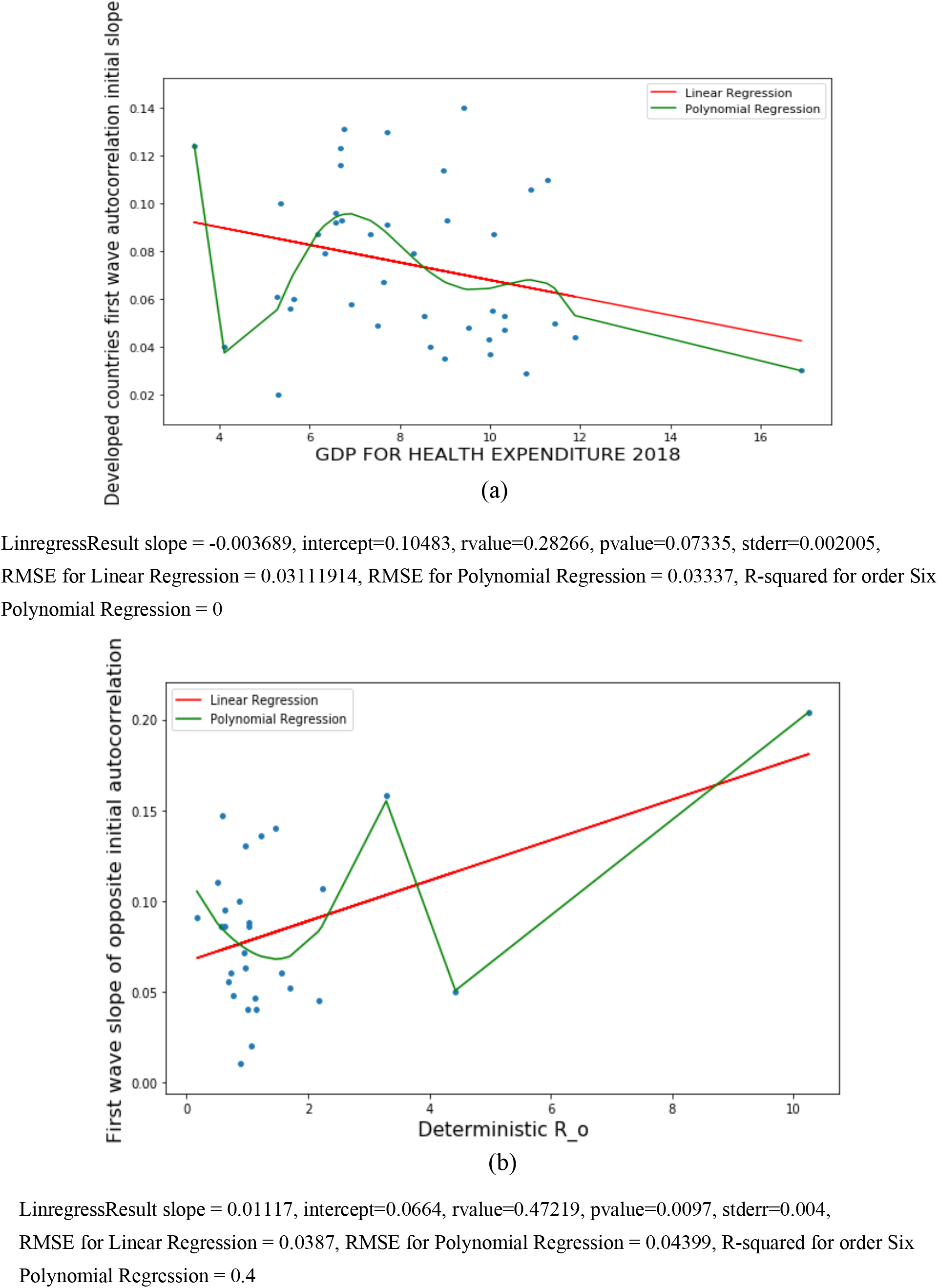
Linear (in red) and sextic (in green) regression plots of first wave opposite of initial autocorrelation slope vs (a) CHE/GDP and (b) maximum R_0_ for developed countries.

### 3.2. Exponential model slope

#### 3.2.1. Developed and Developing countries

**Figure 4.**
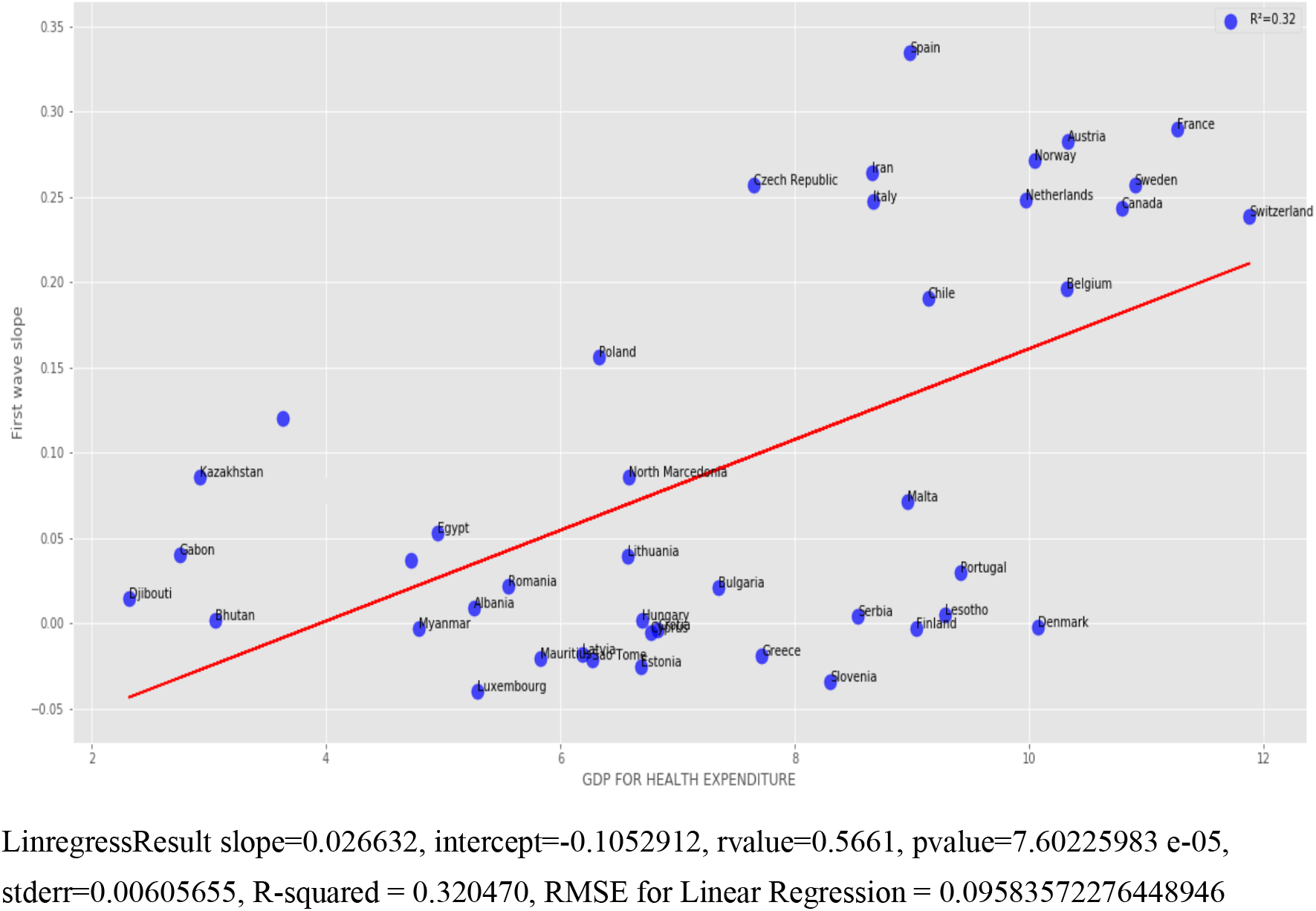
Regression plot of first wave exponential regression slope vs CHE/GDP for developed and developing countries.

#### 3.2.2. Developed countries

**Figure 5.**
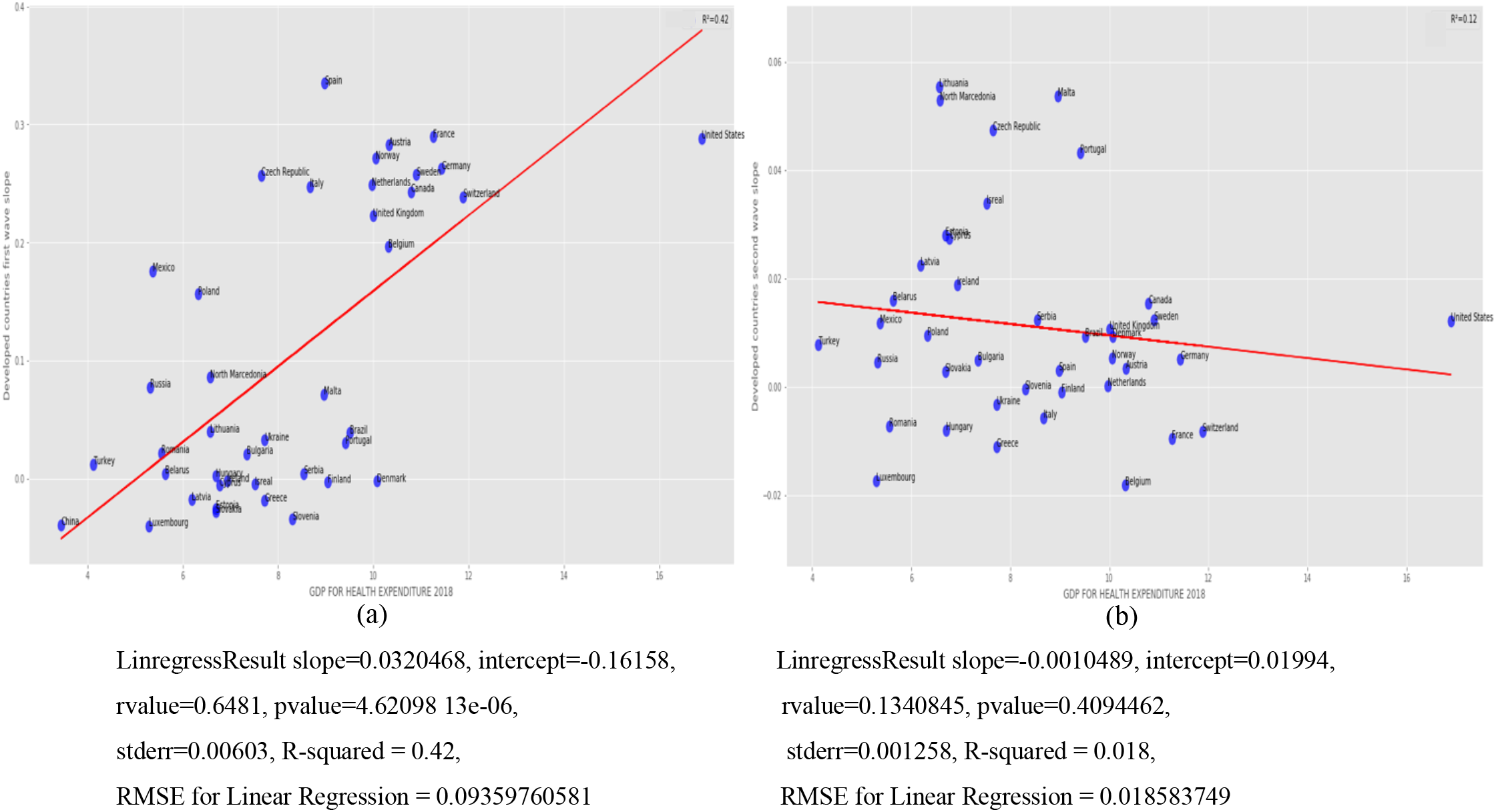
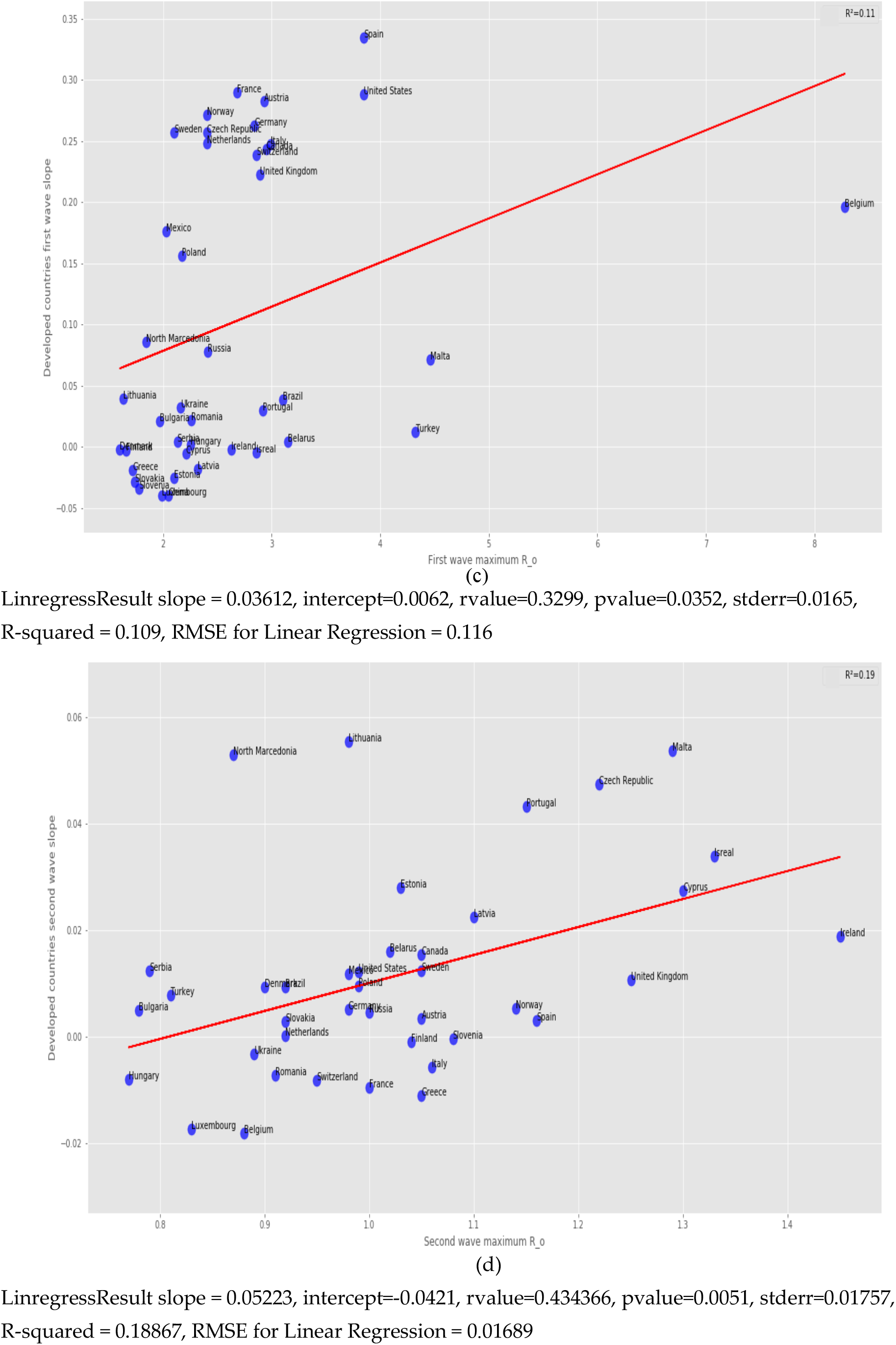
Regression plots for developed countries of (a) first and (b) second wave exponential regression slope versus CHR/GDP, (c) first and (d) second wave maximum R_0_ of the new cases curve.

#### 3.2.3. All countries

**Figure 6.**
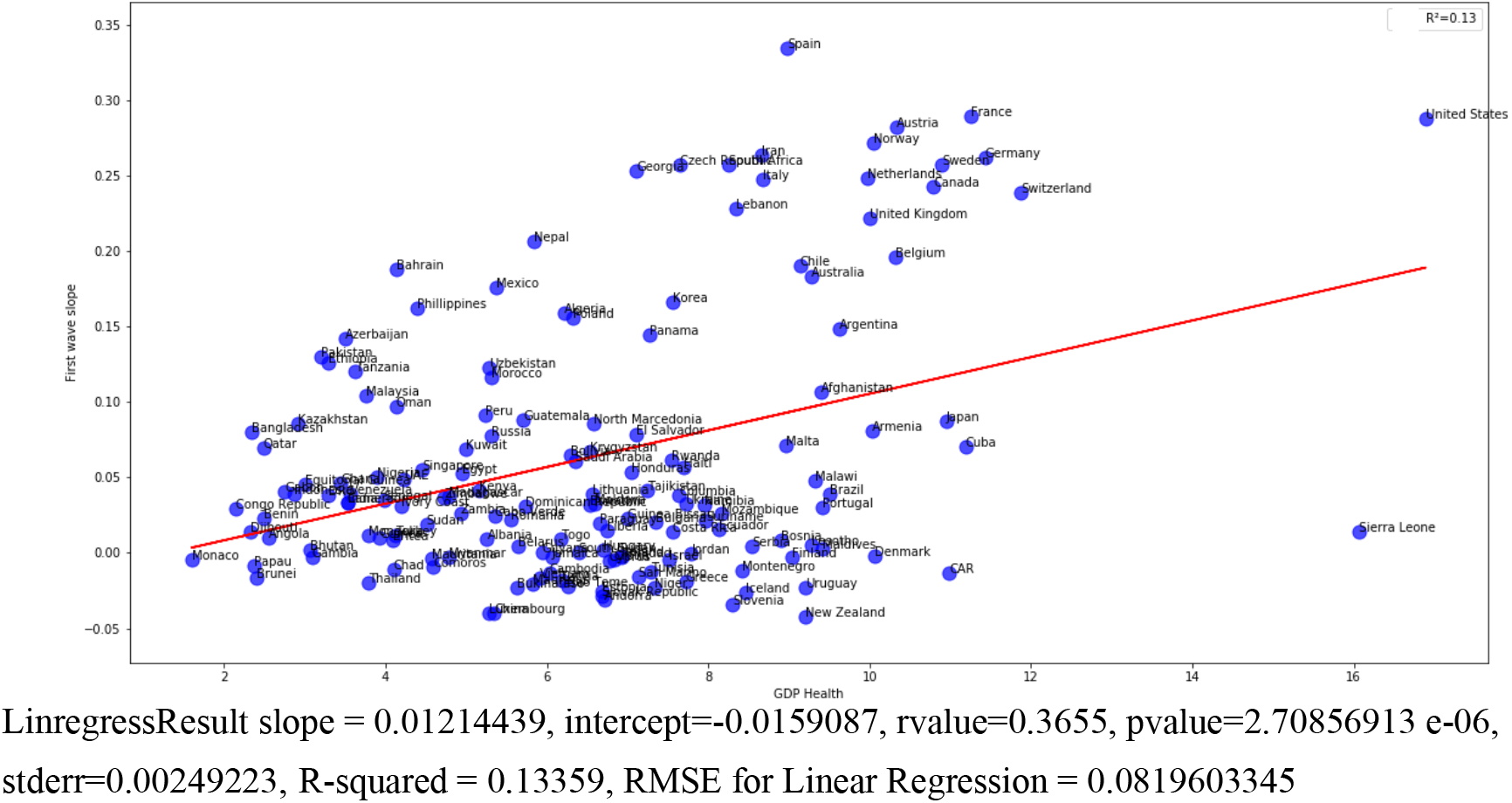
Regression plot of first wave exponential regression slope vs CHE/GDP for all countries.

### 3.3. ARIMA Model for first and second wave

**Figure 7.**
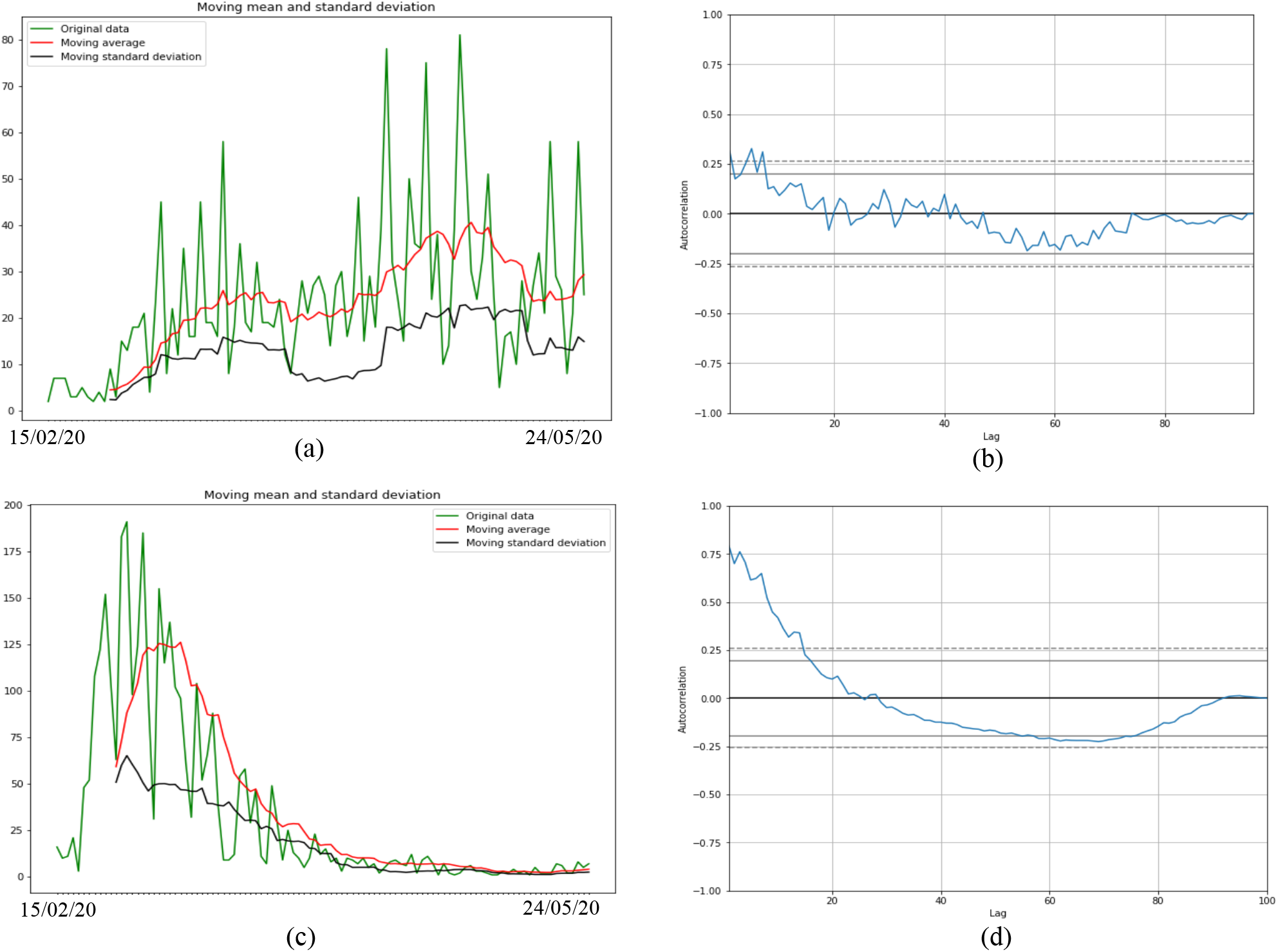
(a) First wave moving average and standard deviation of new cases (left) and (b) autocorrelation curve for Mali (right). (c) First wave moving average and standard deviation of new cases (left) and (d) autocorrelation curve for Luxembourg (right).

#### 3.3.2. Second wave ARIMA Model from October 15 2020 to January 22 2021

**Figure 8.**
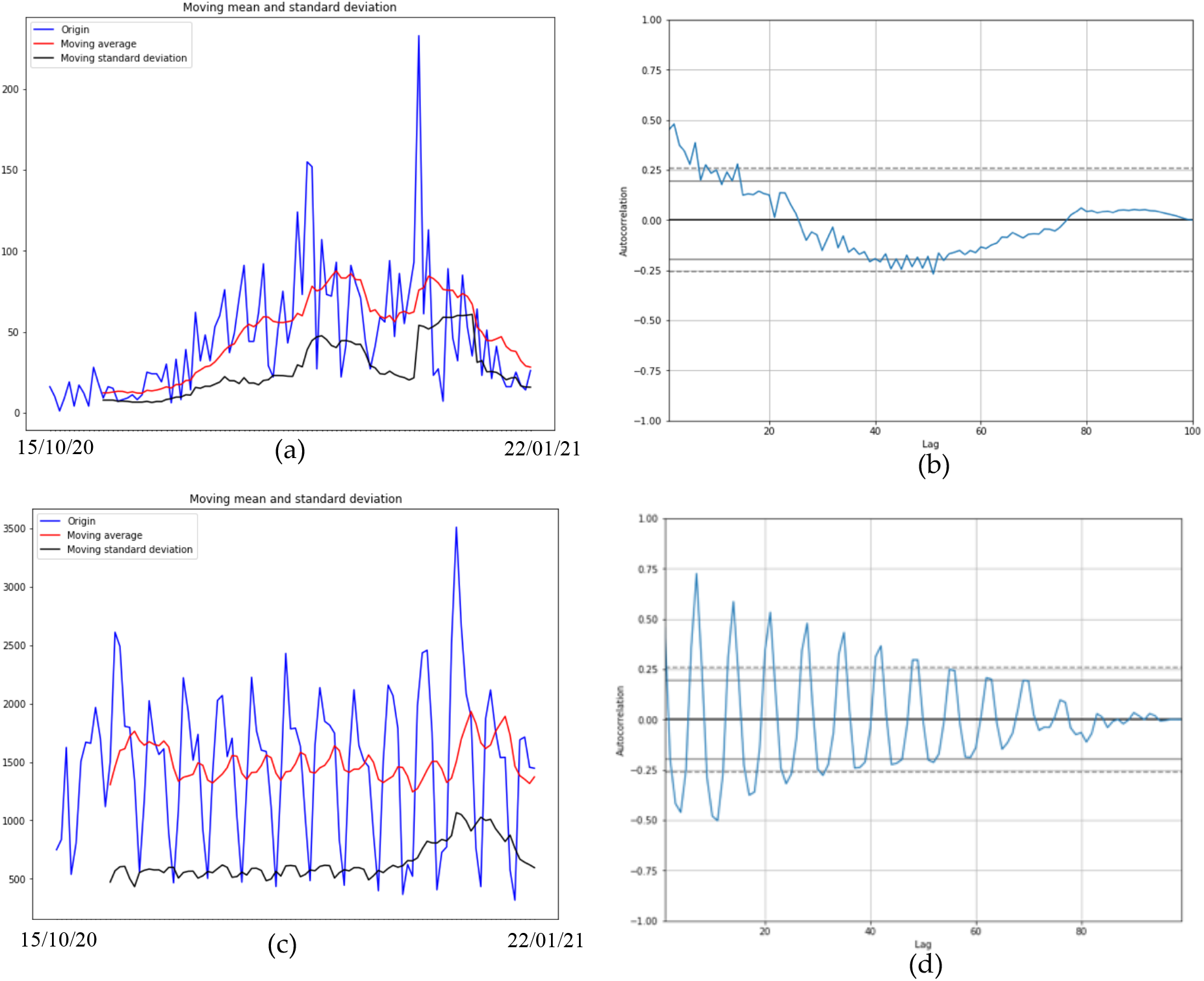
(a) Second wave moving average and standard deviation of new cases (left) and (b) autocorrelation curve for Mali (right). (c) Second wave moving average and standard deviation of new cases (left) and (d) autocorrelation curve for Slovenia (right).

#### 3.3.3. ARIMA Model forecast for first and second wave

**Figure 9.**
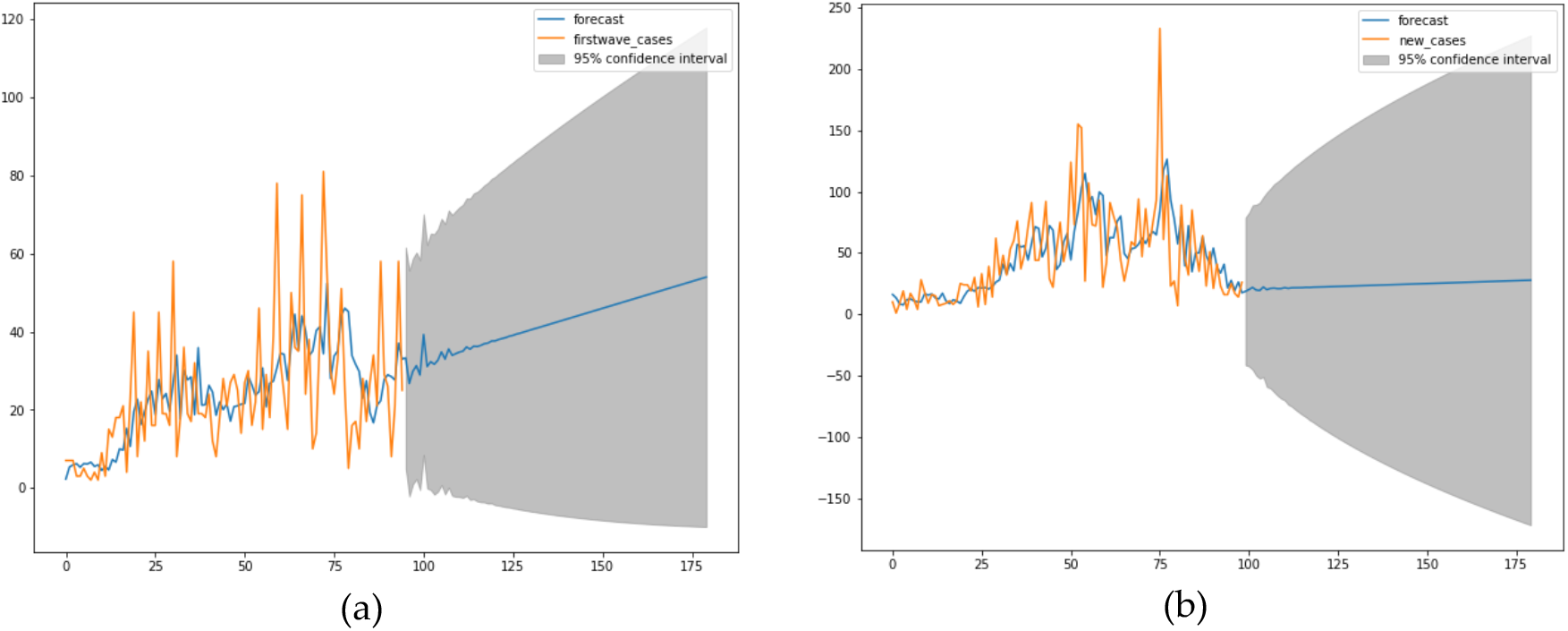
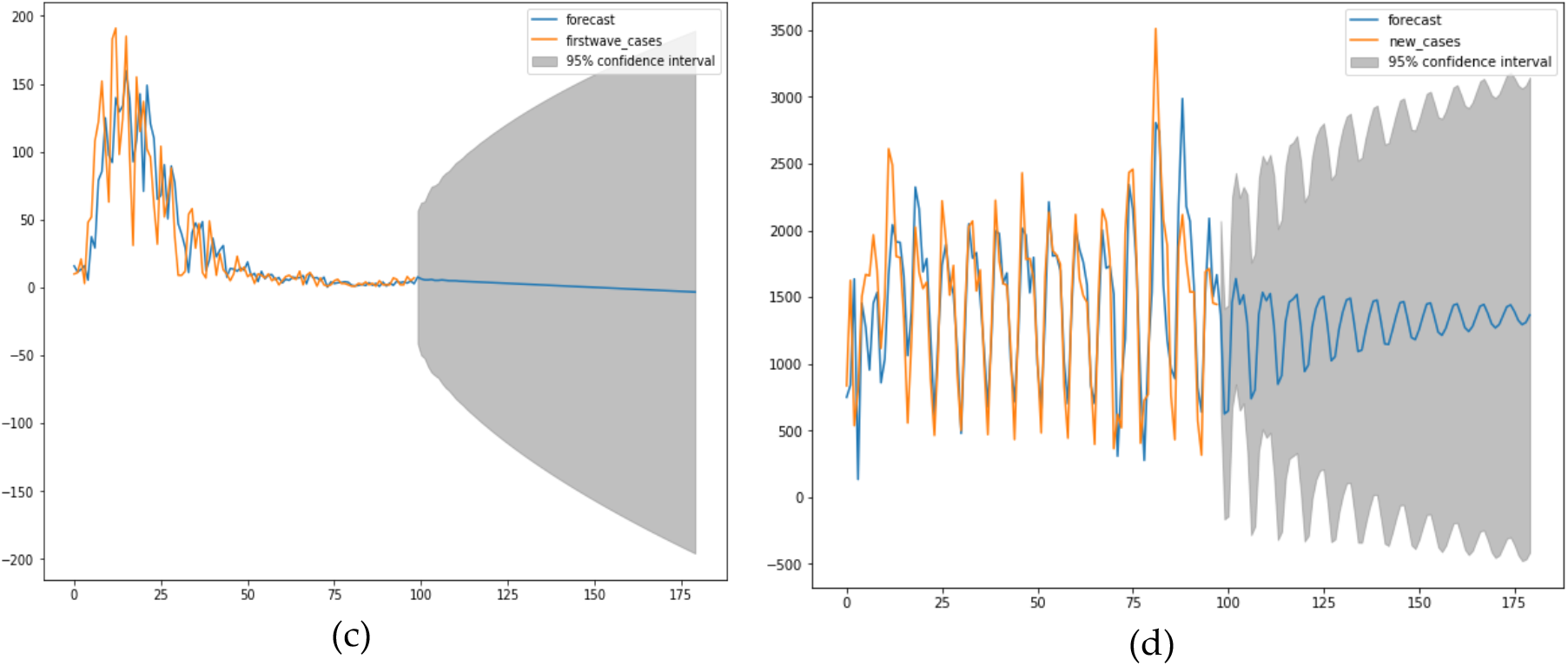
(a) First and (b) second wave forecast for Mali. (c) First wave forecast for Luxembourg. (d) Second wave forecast for Slovenia.

### 3.4. Clustering of variables

#### 3.4.1. Hierarchical clustering

**Figure 10.**
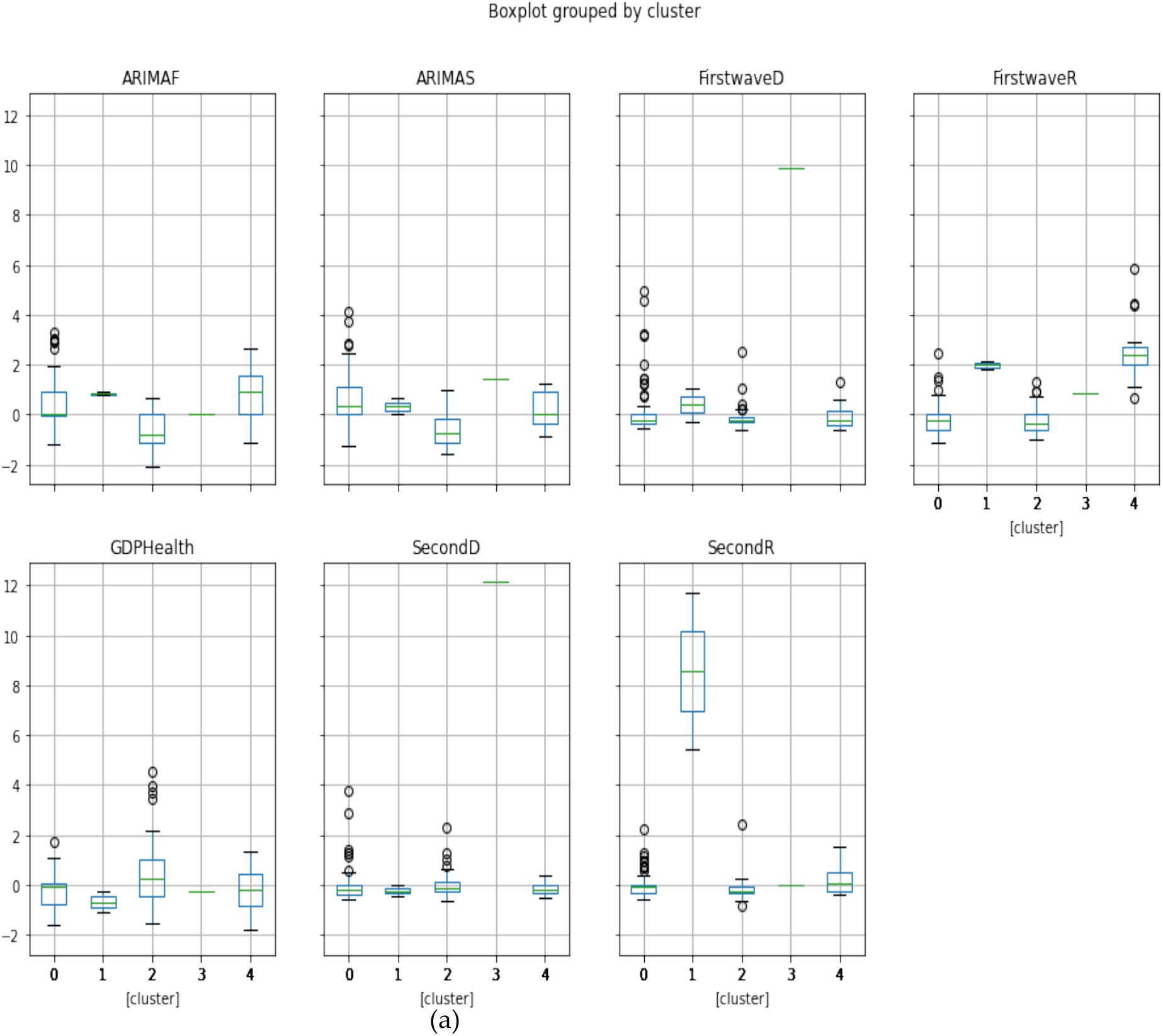
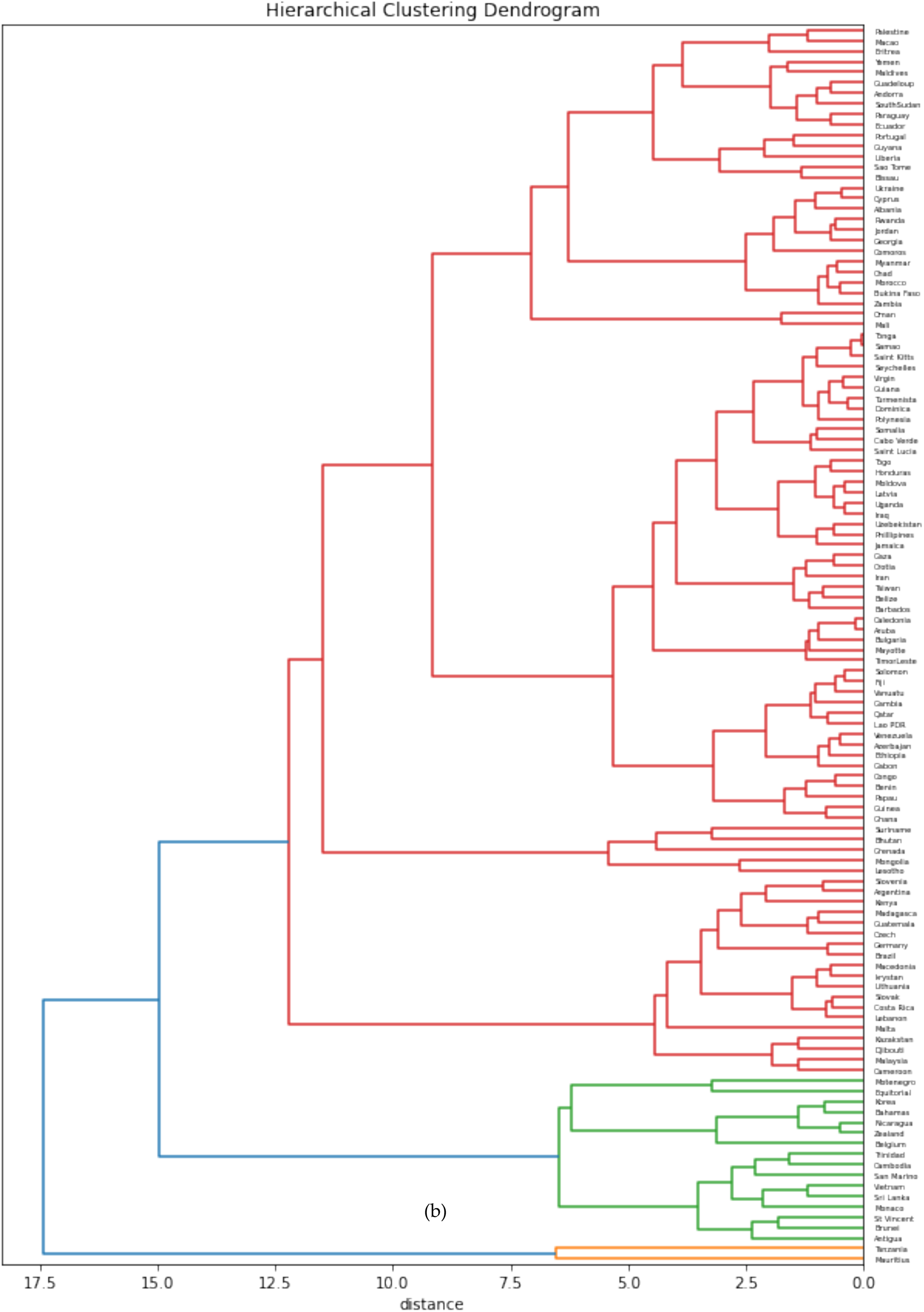
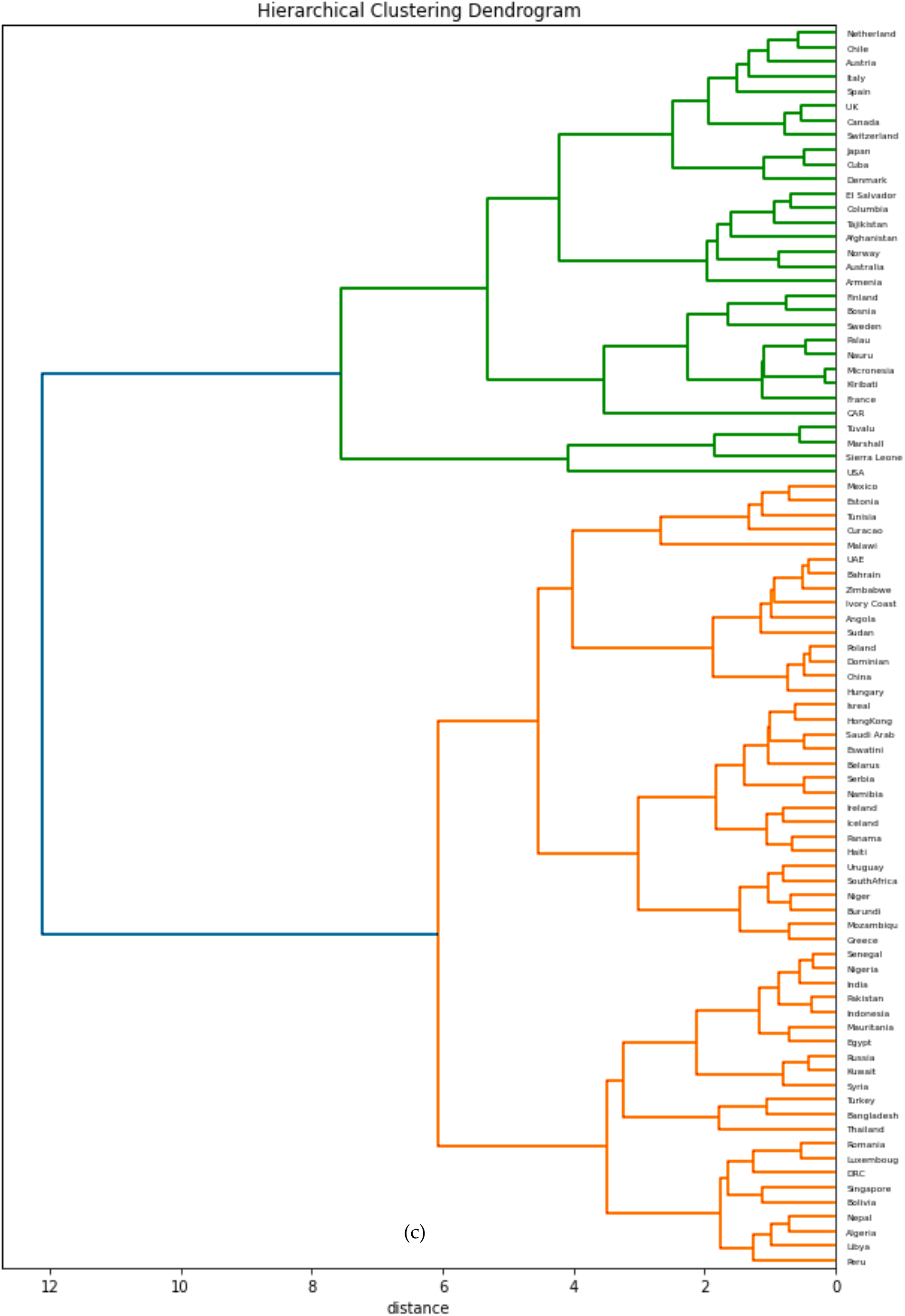
(a) Boxplots of the clusters corresponding to the hierarchical clustering. Visualizations of (b) more “developing” (in red with some notable exceptions like Czech and Germany) and (c) more “developed” (in green and partially in orange) countries parts of the hierarchy tree.

#### 3.4.2. PCA Clustering

**Figure 11.**
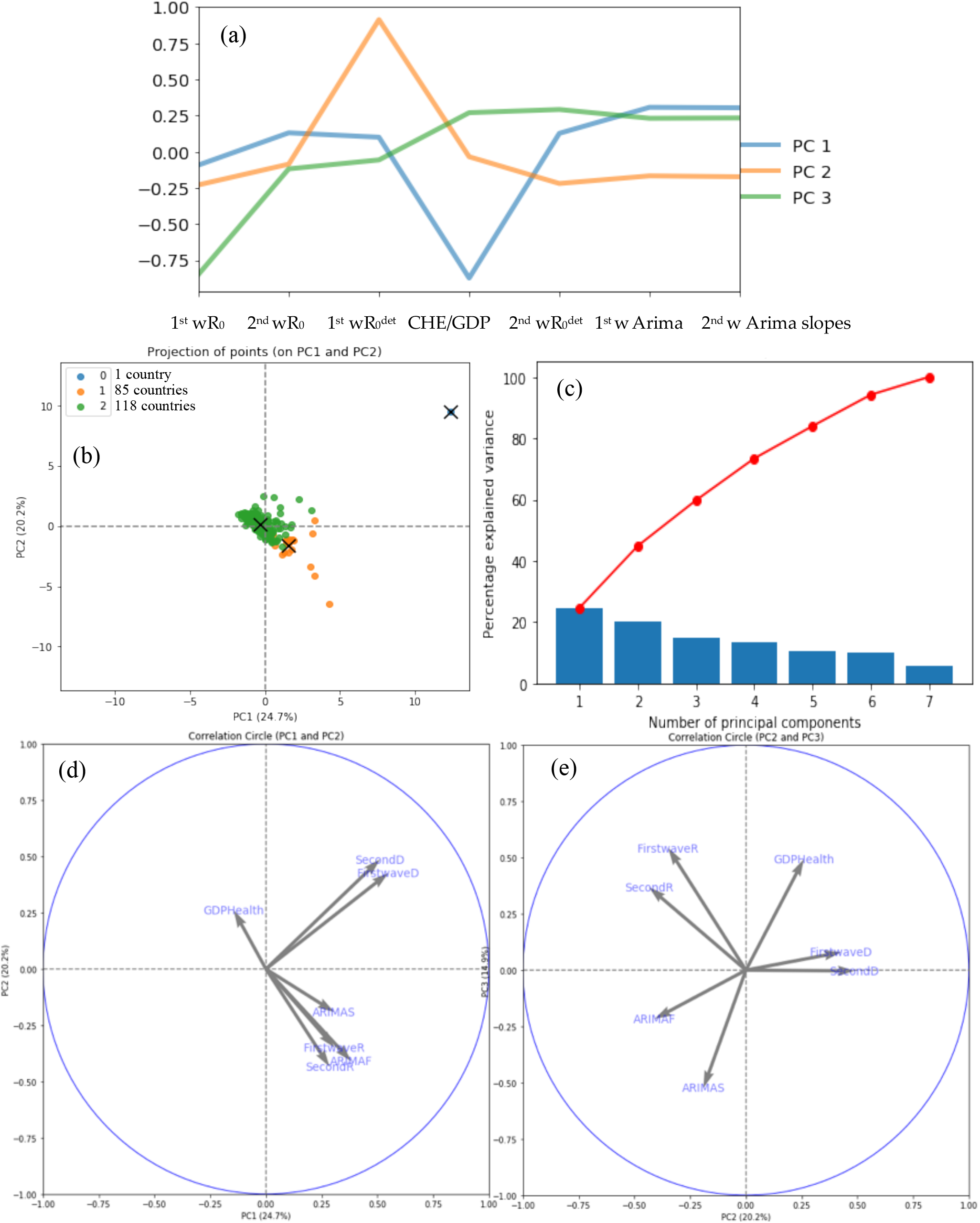
(a) Principal Components (PC) plot from the Principal Component Analysis (PCA) on the initial variables: first and second waves maximum R_0_, 1^st^ wR_0_ and 2^nd^ wR_0_, deterministic R_0_, 1^st^ wR_0_^det^ and 2^nd^ wR_0_^det^, Arima slopes, 1^st^ wArima, 2^nd^ wArima slopes, and CHE/GDP. (b) Projection of the points corresponding to 204 countries of the PCA’s plot on the first PC plane with more developed countries in green and more developing in orange. (c) Explained variance plot. (d&e) Correlation circles for the two first PC planes.

#### 3.4.3. K-means clustering

**Figure 12.**
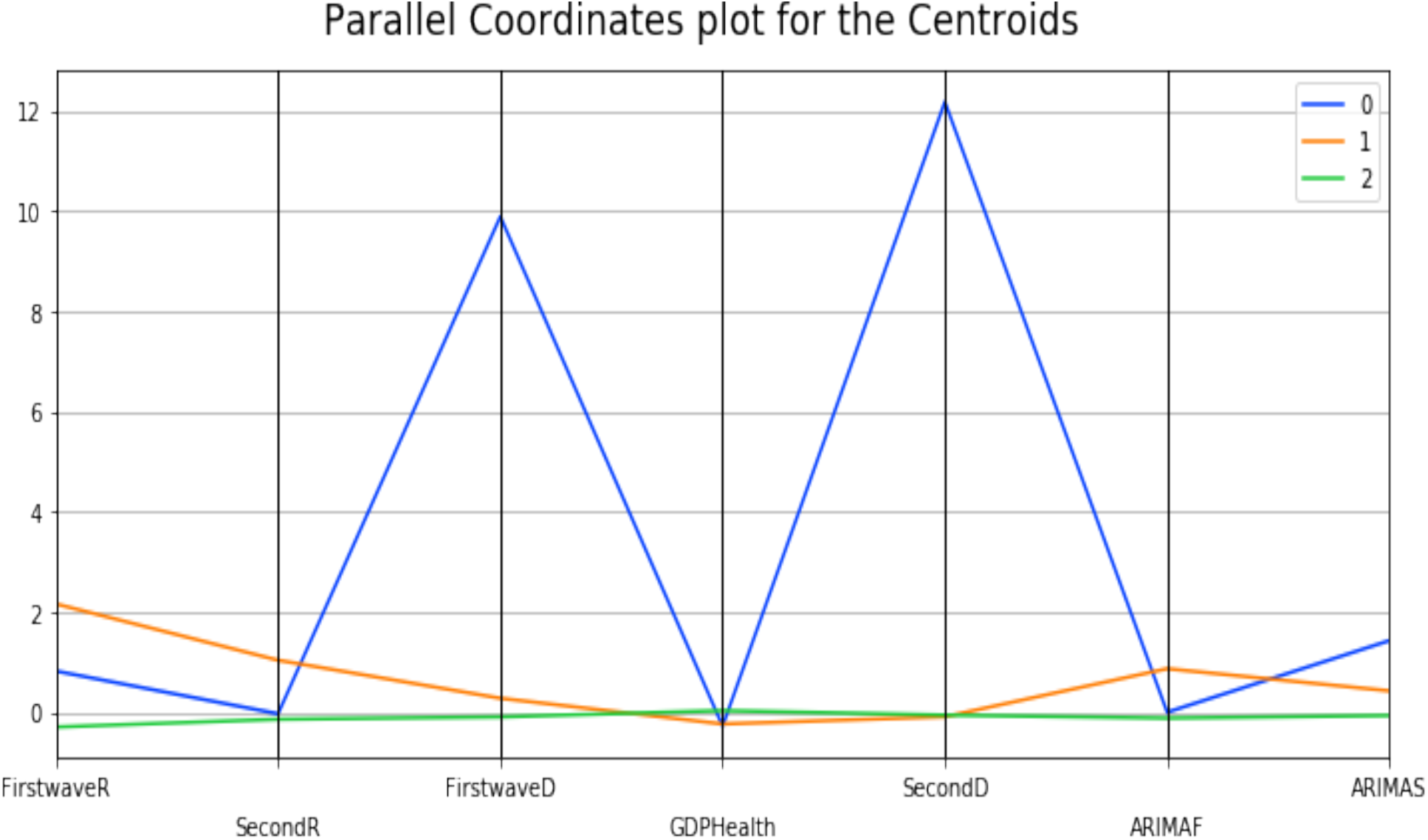
Parallel coordinates for cluster centroids.

### 3.4. Ordinary least square method. The multivariate case

**Figure 13.**
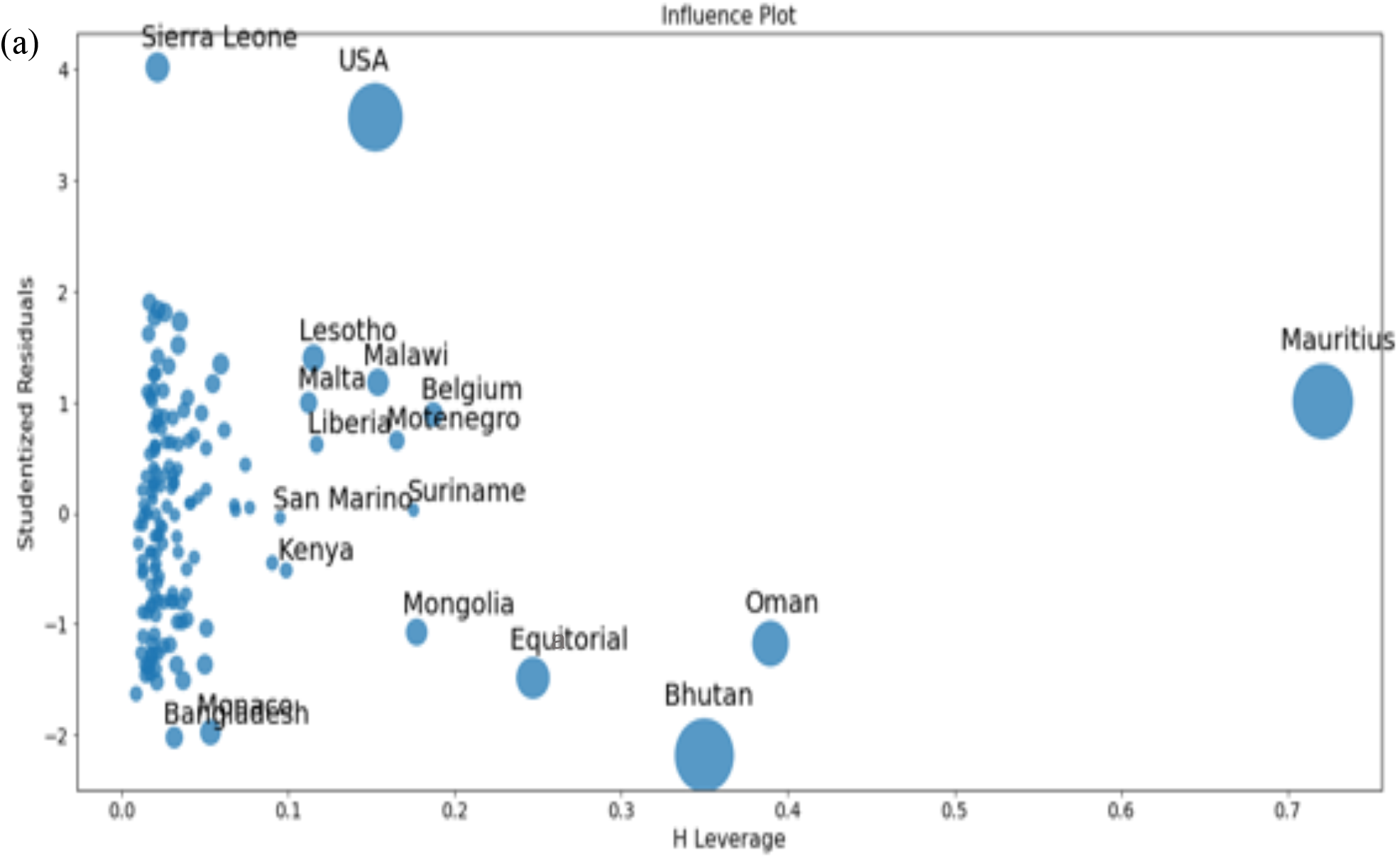
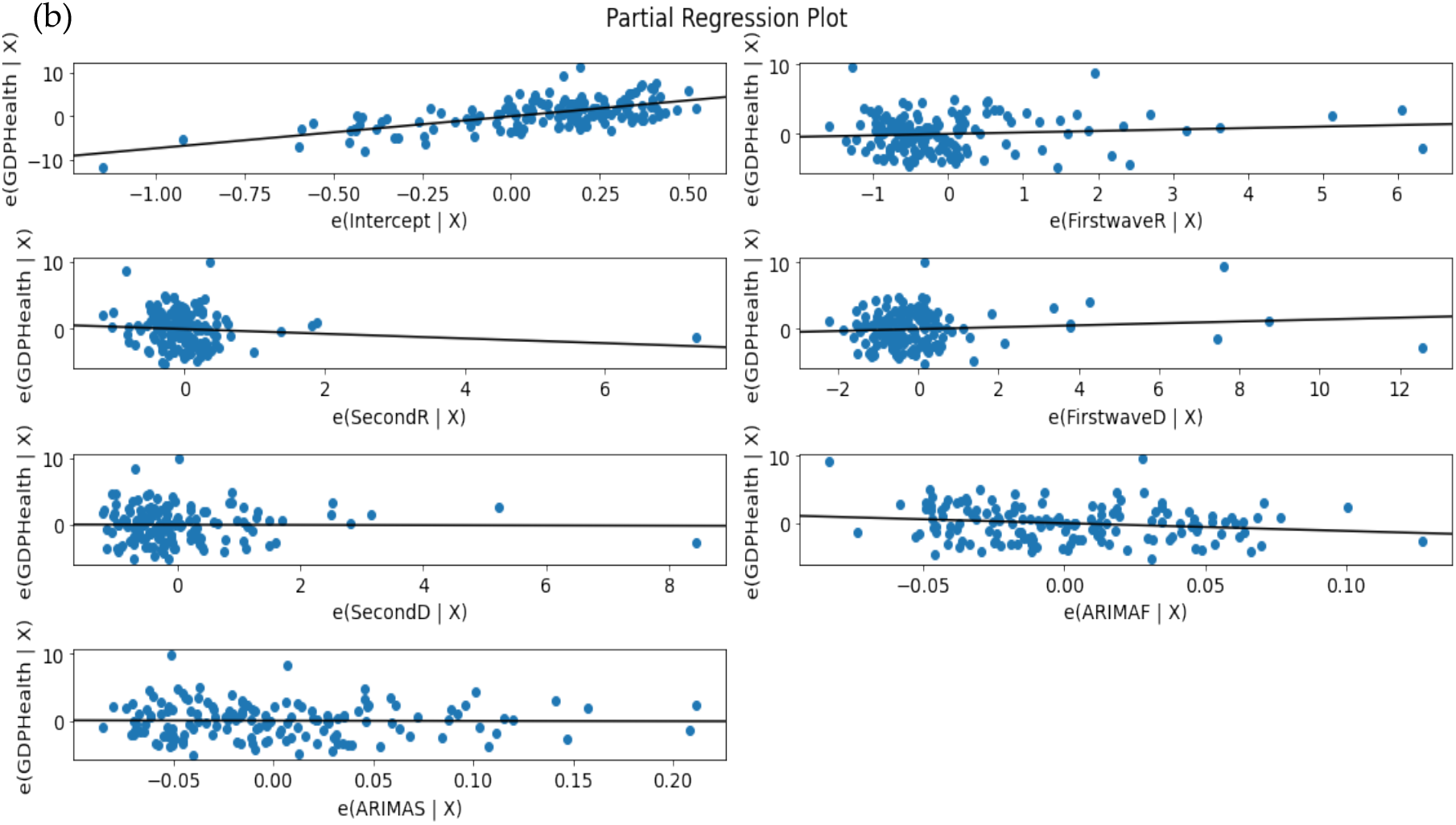
(a) Leverage vs normalized squared residuals plot. (b) Residuals regression plots for initial variables.

## 4. Discussion

There are a lot of differences between the first and second wave results concerning the exponential regression slope and the autocorrelation initial slope: while some countries have higher figures for the first wave, others have lower figures for the second wave and vice versa. This was also evident for the regression plot where some countries have negative correlation values for the first wave of some growth parameters with the CHE/GDP and positive for the second wave, and vice versa for other countries. These phenomena prove that the way the pandemic spread in the second wave is different from what was experienced in the first wave. In the principal component analysis, we discovered that first wave deterministic R_0_ and CHE/GDP health had high weights in first and second Principal Components (PC1 and PC2), which are dominant components in the PC analysis.

More precisely, on Figure 1a,b first and second waves of the Covid-19 pandemic are compared using linear and parabolic or cubic regression, showing a significant positive (resp. negative) correlation between the opposite of the initial autocorrelation slope and exponential regression slope of the first (resp. second) wave for developed (resp. all) countries. This opposition between the two waves could result from the application of a more severe lockdown in developed countries during the second wave. On Figure 1c, the opposite of the initial autocorrelation slope decreases significantly if the start of the first wave in a country is late with respect to the start of the Covid-19 outbreak in China due probably to the progressive implementation of mitigation measures in that country taking into account the experience of the countries starting first wave before. On Figure 2, the opposite of the initial autocorrelation slope is significantly negatively correlated with the maximum R_0_ observed at the inflection point of the new cases curve, confirming that long contagiousness periods give high exponential increases of the new cases. On Figure 3, for the first wave the opposite of the initial autocorrelation slope is positively (resp. negatively) correlated with the CHE/GDP (resp. maximum R_0_) for developed countries, which could correspond to the efficiency of the mitigation measures decided in these countries, which is confirmed on Figure 4, where the first wave exponential regression slope is positively correlated with the CHE/GDP in a mix of developed and developing countries. The Figure 5a shows the same type of effect of public health policies in developed countries for the first wave, where CHE/GDP increases with the first wave exponential regression slope, but this result is inversed on Figure 5b for the second wave perhaps due to a rationalization of the care activity between the first two waves. Figures 5c&d show a similar behavior of the two waves concerning the positive correlation between the exponential regression slope and the maximum R_0_, which makes sense, as these quantities are both related to the initial exponential growth of an epidemic wave. For the first wave of all countries, Figure 6 shows the same positive correlation as Figure 5a between the exponential regression slope and CHE/GDP.

Figure 7 compares two countries, one from sahelian Africa, Mali and one from western Europe, Luxembourg during the first wave of Covid-19 outbreak during the spring 2020: Mali shows a quasi-endemic behavior with a weakly varying autocorrelation function and Luxembourg a frank epidemic wave with a classic shape. For the second wave in fall 2020, Mali presents an attenuated epidemic shape (due probably to specific geoclimatic conditions in western Africa [7]) and a country from central Europe, Slovenia, shows at this period an endemic behavior with an oscillatory occurrence of new cases. Figure 9 proposes a forecasting based on ARIMA decomposition for the first and second waves in Mali with a better approximation for the epidemic second wave than for the quasi-endemic first wave. It is the same for Luxembourg with an inversion of the phases order, an epidemic wave followed by an endemic state well predicted. On the contrary for Slovenia, the endemic state with oscillations is badly predicted.

Clustering of all countries is then studied on Figures 10 to 12. Figure 10a shows the boxplot of the 7 initial variables used in hierarchical clustering: the first and second wave opposite of the initial autocorrelation slope (respectively ARIMAF and ARIMAS), exponential regression slope and maximum R_0_ (respectively FirstwaveD, SecondD, FirstwaveR, SecondR), and the CHE/GDP. The boxplots contain 5 clusters represented in Figure 10b&c corresponding to more “developing” (in red with some notable exceptions like Czech and Germany) and (c) more “developed” (in green and partially in orange) countries parts of the hierarchy tree, with a small “exotic” cluster for Tanzania and Mauritius. Figures 11a-e shows the results of the Principal Component Analysis (PCA), with (a) the 3 principal components declined on the initial variables calculated for all countries (first and second waves maximum R_0_’s denoted 1^st^ wR_0_ and 2^nd^ wR_0_, deterministic R_0_’s denoted 1^st^ wR_0_^det^ and 2^nd^ wR_0_^det^, Arima slopes denoted 1^st^ wArima, 2^nd^ wArima slopes, and the Current Health Expenditure as Gross Domestic Product percentage denoted CHE/GDP), (b) the projection of the points corresponding to countries of the PCA’s plot on the first PC plane, (c) the explained variance plot and (d&e) the correlation circles for the first three Principal Components with projection of the initial variables as vectors (having 195 components corresponding to the 195 countries of the Table 1 in Appendix 1) on the corresponding principal planes. In Figure 11a, the main initial variable in the linear combination of the first (resp. the second) principal component is the first wave deterministic R_0_^det^ (resp. the CHE/GDP) and these two initial variables R_0_^det^ and CHE/GDP are anticorrelated as we have already noticed when commenting before on the Figure 3 (a country devoting a large share of its GDP to health expenditure reduces the occurrence of new cases). Figure 11b gives the projection of 204 countries on the first PC plane and distinguishes 2 main clusters of respectively 118 and 85 countries (plus a singleton representing Botswana), with more developed countries in green and more developing in orange. Figure 11c shows that 60% of the variance is explained by the 3 first PCs, and Figures 11d&e present the correlation circles with projection of the initial variables as vectors on the corresponding two principal planes (PC1, PC2) and (PC2, PC3), showing like in Figure 11a the preeminence of the opposite vectors, the first wave deterministic R_0_ and the CHE/GDP. Figure 12 shows also for the first k-means cluster the importance of the first wave deterministic R_0_.

The last Figures 13a,b correspond to the ordinary multivariate least square method. Figure 13a shows the eccentric position of developed countries like Belgium and USA and developing countries like Equatorial Guinea and Suriname as outliers not fitting the data bulk, and Figure 13b the concentration of the initial variable CHE/GDP with the first and second waves deterministic R_0_^det^, in agreement with the fact that they are the most dominant initial variables in PCA and k-means clustering.

## 5. Conclusions

We have shown in this article that there exist correlations between the growth parameters directly linked to the occurrence of new cases of Covid-19 and socio-economic variables, in particular the Current Health Expenditure as Gross Domestic Product percentage (CHE/GDP) anticorrelated with the basic reproduction time R_0_, which shows the effectiveness of public health mitigation measures, even if they involve significant medico-economic costs. Larger perspectives are offered by combining other studies on geoclimatic and demographic factors of severity of the Covid-19 outbreak [7-13] with the present socio-economic determinants, in order to get the most comprehensive and accurate picture of non-biological exogenous influences on the expanding Covid-19 pandemic.

## Data Availability

All the data are coming from public databases

## Appendix A

**Table 1.** Parameters for first and second waves for 195 countries.

| All countries |  |  |  |  |  |  |  |  |  |  |
| --- | --- | --- | --- | --- | --- | --- | --- | --- | --- | --- |
| First wave |  |  |  |  |  | Second wave |  |  |  |  |
| No | Country Name | $R_0$ | D-$R_0$ | Exponential Slope | Auto-correlation slope | $R_0$ | D-$R_0$ | Exponential Slope | Auto-correlation slope | GDP Health 2018 |
| 1 | AFGHANISTAN | 1.78 | 0.65 | 0.1070 | -0.025 | 0.79 | 0.04 | 0.0017 | -0.097 | 9.40 |
| 2 | ALGERIA | 2.19 | 1.25 | 0.1594 | -0.040 | 0.86 | 0.91 | 0.0316 | -0.100 | 6.22 |
| 3 | ARUBA | - | 5.46 | - | - | - | 1.10 | 0.0172 | -0.112 | - |
| 4 | ANDORRA | - | 1.36 | -0.0313 | -0.121 | - | 0.12 | -0.0067 | -0.155 | 6.71 |
| 5 | ANGOLA | 2.06 | 0.63 | 0.0100 | -0.095 | 1.13 | 1.70 | -0.0135 | -0.057 | 2.55 |
| 6 | ANTIGUA | 4.23 | 1.92 | - | - | 3.30 | 2.13 | 0.0051 | -0.177 | 5.23 |
| 7 | ALBANIA | 1.61 | 0.96 | 0.0091 | -0.138 | 0.99 | 0.66 | 0.0058 | -0.085 | 5.26 |
| 8 | ARGENTINA | 2.06 | 0.73 | 0.1485 | -0.060 | 1.19 | 0.36 | 0.0427 | -0.240 | 9.62 |
| 9 | ARMENIA | 1.51 | 4.43 | 0.0809 | -0.050 | 0.80 | 0.86 | 0.0570 | -0.090 | 10.03 |
| 10 | AUSTRALIA | 2.45 | 2.79 | 0.1832 | -0.054 | 1.11 | 1.50 | 0.0037 | -0.136 | 9.28 |
| 11 | AUSTRIA | 2.93 | 1.17 | 0.2825 | -0.053 | 1.05 | 2.08 | 0.0034 | -0.053 | 10.33 |
| 12 | AZERBAIJAN | 2.11 | 1.16 | 0.1422 | -0.071 | 0.63 | 0.37 | 0.0676 | -0.130 | 3.51 |
| 13 | BAHAMAS | 6.33 | 0.57 | - | - | 1.48 | 1.22 | -0.0250 | -0.077 | 6.25 |
| 14 | BAHRAIN | 1.81 | 1.10 | 0.1884 | -0.079 | 1.24 | 1.14 | 0.0012 | -0.053 | 4.13 |
| 15 | BANGLADESH | 3.67 | 1.04 | 0.0799 | -0.033 | 0.92 | 0.99 | -0.0086 | -0.046 | 2.34 |
| 16 | BARBADOS | 4.63 | 1.86 | - | - | 1.99 | 1.14 | 0.0378 | -0.109 | 6.56 |
| 17 | BELARUS | 3.15 | 1.57 | 0.0043 | -0.060 | 1.02 | 1.07 | 0.0159 | -0.026 | 5.64 |
| 18 | BELGIUM | 8.28 | 0.43 | 0.1963 | -0.047 | 0.88 | 2.23 | -0.0182 | -0.063 | 10.32 |
| 19 | BELIZE | 3.74 | 0.99 | - | - | 1.34 | 0.51 | -0.0004 | -0.140 | 5.69 |
| 20 | BENIN | 2.16 | 0.85 | 0.0226 | -0.133 | 1.55 | 0.85 | 0.0020 | -0.125 | 2.49 |
| 21 | BHUTAN | 2.10 | 15.00 | 0.0021 | -0.118 | 2.49 | 1.08 | 0.0126 | -0.099 | 3.06 |
| 22 | BOLIVIA | 1.46 | 2.17 | 0.0647 | -0.045 | 1.45 | 1.61 | 0.0152 | -0.087 | 6.30 |
| 23 | BOSNIA | 1.70 | 0.09 | 0.0088 | -0.110 | 0.97 | 1.56 | -0.0118 | -0.106 | 8.90 |
| 24 | BOTSWANA | 3.76 | 28.47 | - | - | 1.43 | 28.43 | 0.0030 | -0.186 | 5.85 |
| 25 | BRAZIL | 3.10 | 0.77 | 0.0389 | -0.048 | 0.92 | 0.46 | 0.0092 | -0.188 | 9.51 |
| 26 | BRUNEI | 5.00 | 1.08 | -0.0165 | -0.120 | 3.66 | 1.00 | - | - | 2.41 |
| 27 | BULGARIA | 1.97 | 5.06 | 0.0178 | -0.087 | 0.78 | 0.75 | 0.0049 | -0.110 | 7.35 |
| 28 | BURKINA FASO | 2.44 | 1.08 | -0.0227 | -0.123 | 1.18 | 0.94 | 0.0360 | -0.058 | 5.63 |
| 29 | BURUNDI | 2.80 | 1.33 | - | - | 1.69 | 2.18 | 0.0226 | -0.063 | 7.74 |
| 30 | CABO VERDE | 1.54 | 0.82 | 0.0247 | -0.091 | 1.71 | 0.19 | -0.0064 | -0.110 | 5.36 |
| 31 | CAMBODIA | 5.55 | 0.34 | -0.0129 | -0.129 | 3.12 | 0.27 | 0.0010 | -0.158 | 6.03 |
| 32 | CAMEROON | 2.56 | 2.17 | 0.0338 | -0.123 | 1.64 | 2.48 | 0.0085 | -0.207 | - |
| 33 | CANADA | 2.95 | 1.10 | 0.2432 | -0.029 | 1.05 | 0.44 | 0.0153 | -0.047 | 10.79 |
| 34 | CAR | 2.45 | 1.66 | -0.0130 | -0.096 | 4.99 | 0.33 | - | - | 10.99 |
| 35 | CHAD | 2.43 | 1.19 | -0.0108 | -0.114 | 1.44 | 0.77 | 0.0222 | -0.050 | 4.10 |
| 36 | CHILE | 2.42 | 1.00 | 0.1906 | -0.034 | 1.16 | 1.64 | 0.0586 | -0.090 | 9.14 |
| 37 | CHINA | 2.05 | 1.10 | -0.0602 | -0.088 | 1.07 | 0.87 | 0.0137 | -0.068 | 5.35 |
| 38 | COLUMBIA | 1.86 | 1.00 | 0.0384 | -0.040 | 0.99 | 1.47 | 0.0061 | -0.126 | 7.64 |
| 39 | COMOROS | 1.93 | 3.75 | -0.0094 | -0.153 | 1.58 | 1.65 | 0.0397 | -0.076 | 4.59 |
| 40 | CONGO DEM | 1.48 | 0.03 | 0.0384 | -0.052 | 1.10 | 0.88 | 0.0252 | -0.089 | 3.30 |
| 41 | CONGO REP | 2.39 | 0.92 | 0.0294 | -0.152 | 1.43 | 0.39 | 0.0064 | -0.118 | 2.14 |
| 42 | COSTA RICA | 1.51 | 0.50 | 0.0142 | -0.110 | 1.08 | 1.26 | -0.0022 | -0.209 | 7.56 |
| 43 | COTE D'VOIRE | 1.47 | 1.18 | 0.0309 | -0.080 | 1.35 | 2.09 | 0.0253 | -0.078 | 4.19 |
| 44 | CROTIA | 3.95 | 0.75 | -0.0042 | -0.069 | 0.72 | 0.57 | -0.0115 | -0.106 | 6.83 |
| 45 | CUBA | 2.23 | 0.48 | 0.0706 | -0.063 | 1.30 | 0.78 | 0.0517 | -0.040 | 11.19 |
| 46 | CURACAO | - | 0.50 | - | - | - | 4.19 | -0.0060 | -0.074 | - |
| 47 | CYPRUS | 2.21 | 0.69 | -0.0056 | -0.131 | 1.30 | 0.45 | 0.0273 | -0.089 | 6.77 |
| 48 | CZECH | 2.40 | 0.16 | 0.2570 | -0.067 | 1.22 | 0.88 | 0.0474 | -0.197 | 7.65 |
| 49 | DENMARK | 1.60 | 0.80 | -0.0024 | -0.087 | 0.90 | 0.64 | 0.0092 | -0.048 | 10.07 |
| 50 | DJIBOUTI | 2.73 | 0.17 | 0.0144 | -0.094 | 1.47 | 0.36 | -0.0045 | -0.169 | 2.32 |
| 51 | DOMINICAN | 2.09 | 1.02 | 0.0309 | -0.088 | 1.10 | 1.57 | 0.0151 | -0.081 | 5.73 |
| 52 | DOMINICA | - | 7.75 | - | - | - | 0.67 | - | - | 6.59 |
| 53 | ECUADOR | 2.22 | 1.46 | 0.0157 | -0.140 | 1.18 | 1.14 | -0.0045 | -0.175 | 8.14 |
| 54 | EGYPT | 1.69 | 0.84 | 0.0527 | -0.042 | 1.33 | 0.51 | 0.0243 | -0.023 | 4.95 |
| 55 | EL SALVADOR | 1.58 | 1.70 | 0.0783 | -0.052 | 1.29 | 0.66 | 0.0535 | -0.113 | 7.11 |
| 56 | EQUATORIAL G. | 10.0 | 0.38 | 0.0454 | -0.190 | 2.41 | 1.48 | 0.0142 | -0.177 | 3.00 |
| 57 | ERITREA | 2.57 | 1.18 | 0.0083 | -0.216 | 0.74 | 0.80 | 0.0222 | -0.146 | 4.09 |
| 58 | ESTONIA | 2.10 | 0.87 | -0.0254 | -0.116 | 1.03 | 3.04 | 0.0279 | -0.099 | 6.69 |
| 59 | ESWATINI | 2.08 | 0.94 | 0.0317 | -0.071 | 1.34 | 0.71 | 0.0412 | -0.034 | 6.54 |
| 60 | ETHIOPIA | 2.42 | 0.80 | 0.1259 | -0.054 | 1.11 | 1.24 | -0.0041 | -0.136 | 3.30 |
| 61 | FIJI | - | 2.00 | - | - | - | 0.50 | - | - | 3.42 |
| 62 | FINLAND | 1.66 | 1.14 | -0.0030 | -0.093 | 1.04 | 2.41 | -0.0010 | -0.119 | 9.04 |
| 63 | FRANCE | 2.68 | 1.17 | 0.2898 | -0.110 | 1.00 | 2.17 | -0.0096 | -0.081 | 11.26 |
| 64 | GABON | 1.83 | 0.97 | 0.0404 | -0.077 | 1.44 | 0.19 | 0.0187 | -0.143 | 2.75 |
| 65 | GAMBIA | 3.21 | 0.83 | -0.0026 | -0.094 | 2.29 | 0.37 | 0.0145 | -0.099 | 3.09 |
| 66 | GEORGIA | 2.19 | 1.23 | 0.2536 | -0.136 | 0.76 | 0.79 | 0.0293 | -0.057 | 7.11 |
| 67 | GERMANY | 2.84 | 0.73 | 0.2624 | -0.050 | 0.98 | 0.79 | 0.0050 | -0.195 | 11.43 |
| 68 | GHANA | 1.85 | 1.48 | 0.0463 | -0.099 | 1.09 | 0.62 | 0.0118 | -0.117 | 3.45 |
| 69 | GREECE | 1.72 | 0.71 | -0.0189 | -0.091 | 1.05 | 0.71 | -0.0111 | -0.069 | 7.72 |
| 70 | GRENADA | 5.78 | 14.00 | - | - | 1.08 | 0.10 | 0.0106 | -0.167 | 4.46 |
| 71 | GUADELOUPE | - | 1.35 | -0.0131 | -0.130 | - | 1.35 | -0.0084 | -0.137 | - |
| 72 | GUATEMALA | 1.67 | 0.25 | 0.0880 | -0.044 | 1.08 | 0.27 | 0.1109 | -0.197 | 5.71 |
| 73 | GUIANA FRENCH | - | 0.88 | 0.0391 | -0.102 | - | 0.43 | 0.0238 | -0.124 | - |
| 74 | GUINEA | 1.50 | 0.46 | 0.0097 | -0.111 | 1.36 | 1.68 | -0.0108 | -0.126 | 3.93 |
| 75 | GUINEA BISSAU | 3.56 | 1.14 | 0.0230 | -0.145 | 4.66 | 4.20 | - | - | 7.00 |
| 76 | GUYANA | 2.49 | 2.38 | 0.0005 | -0.152 | 1.54 | 4.23 | -0.0021 | -0.163 | 5.94 |
| 77 | HAITI | 2.32 | 0.60 | 0.0565 | -0.047 | 1.66 | 0.61 | 0.0217 | -0.082 | 7.69 |
| 78 | HONDURAS | 1.96 | 0.57 | 0.0532 | -0.086 | 1.59 | 1.64 | 0.0016 | -0.141 | 7.05 |
| 79 | HONGKONG | - | 0.04 | -0.0003 | -0.060 | - | 0.24 | 0.0285 | -0.041 | - |
| 80 | HUNGARY | 2.25 | 0.90 | 0.0018 | -0.093 | 0.77 | 1.93 | -0.0081 | -0.088 | 6.70 |
| 81 | ICELAND | 2.89 | 2.28 | -0.0261 | -0.056 | 1.86 | 0.66 | -0.0174 | -0.079 | 8.47 |
| 82 | INDIA | 2.43 | 0.98 | 0.0331 | -0.050 | 0.91 | 0.96 | -0.0151 | -0.048 | 3.54 |
| 83 | INDONESIA | 2.04 | 0.95 | 0.0391 | -0.071 | 1.07 | 0.99 | 0.0127 | -0.051 | 2.87 |
| 84 | IRAN | 3.61 | 1.04 | 0.2641 | -0.063 | 1.00 | 0.90 | 0.0438 | -0.140 | 8.66 |
| 85 | IRAQ | 1.81 | 0.77 | 0.1184 | -0.084 | 0.96 | 0.96 | 0.0410 | -0.150 | - |
| 86 | IRELAND | 2.63 | 2.16 | -0.0021 | -0.058 | 1.45 | 1.12 | 0.0188 | -0.057 | 6.93 |
| 87 | ISRAEL | 2.86 | 0.21 | -0.0047 | -0.049 | 1.33 | 1.16 | 0.0339 | -0.037 | 7.52 |
| 88 | ITALY | 2.99 | 1.04 | 0.2475 | -0.040 | 1.06 | 3.69 | -0.0057 | -0.072 | 8.67 |
| 89 | JAMAICA | 2.43 | 0.43 | -0.0031 | -0.089 | 1.22 | 2.47 | 0.0034 | -0.174 | 6.06 |
| 90 | JAPAN | 1.91 | 1.02 | 0.0872 | -0.055 | 1.21 | 1.16 | 0.0260 | -0.052 | 10.95 |
| 91 | JORDAN | 2.16 | 2.53 | -0.0006 | -0.155 | 0.93 | 0.93 | -0.0138 | -0.053 | 7.79 |
| 92 | KAZAKHSTAN | 2.85 | 0.60 | 0.0856 | -0.064 | 1.05 | 2.06 | 0.0933 | -0.210 | 2.92 |
| 93 | KENYA | 1.57 | 1.14 | 0.0413 | -0.067 | 1.26 | 1.18 | -0.0237 | -0.310 | 5.17 |
| 94 | KOREA REP. | 6.06 | 1.00 | 0.1664 | -0.076 | 0.90 | 1.04 | 0.0585 | -0.090 | 7.56 |
| 95 | KOSOVO | 1.90 | 1.02 | - | - | 0.82 | 0.99 | - | - | - |
| 96 | KUWAIT | 2.25 | 0.88 | 0.0687 | -0.031 | 1.27 | 1.10 | -0.0094 | -0.038 | 5.00 |
| 97 | KYRGYZSTAN | 2.27 | 0.17 | 0.0671 | -0.091 | 0.86 | 1.05 | 0.0271 | -0.200 | 6.53 |
| 98 | LAO PDR | - | 0.50 | - | - | - | 0.15 | - | - | 2.25 |
| 99 | LATVIA | 2.32 | 0.74 | -0.0179 | -0.087 | 1.10 | 0.50 | 0.0224 | -0.136 | 6.19 |
| 100 | LEBANON | 1.91 | 1.03 | 0.2286 | -0.112 | 1.27 | 0.90 | 0.0757 | -0.180 | 8.35 |
| 101 | LESOTHO | 1.99 | 7.08 | 0.0053 | -0.206 | 1.36 | 1.42 | 0.0398 | -0.087 | 9.28 |
| 102 | LIBERIA | 1.76 | 0.31 | 0.0151 | -0.114 | 3.08 | 4.56 | 0.0046 | -0.159 | 6.74 |
| 103 | LIBYA | 3.12 | 0.96 | 0.0493 | -0.047 | 1.09 | 0.79 | -0.0059 | -0.099 | - |
| 104 | LITHUANIA | 1.63 | 0.83 | 0.0394 | -0.096 | 0.98 | 2.49 | 0.0554 | -0.230 | 6.57 |
| 105 | LUXEMBOURG | 1.99 | 0.24 | -0.0401 | -0.061 | 0.83 | 1.48 | -0.0174 | -0.105 | 5.29 |
| 106 | MACAO | - | 0.29 | -0.0019 | -0.190 | - |  | - | - | - |
| 107 | MADAGASCAR | 2.48 | 0.94 | 0.0377 | -0.057 | 1.54 | 0.75 | 0.0060 | -0.211 | 4.79 |
| 108 | MALAWI | 3.55 | 1.12 | 0.0478 | -0.088 | 1.66 | 6.46 | 0.0583 | -0.087 | 9.33 |
| 109 | MALAYSIA | 2.86 | 1.25 | 0.1042 | -0.101 | 1.15 | 1.30 | 0.0794 | -0.260 | 3.76 |
| 110 | MALDIVES | 1.96 | 0.83 | 0.0031 | -0.154 | 1.41 | 1.05 | 0.0007 | -0.116 | 9.41 |
| 111 | MALI | 1.61 | 0.64 | 0.0158 | -0.100 | 0.97 | 7.78 | 0.0148 | -0.115 | - |
| 112 | MALTA | 4.46 | 1.06 | 0.0712 | -0.114 | 1.29 | 0.99 | 0.0536 | -0.330 | 8.96 |
| 113 | MAURITANIA | 1.66 | 1.76 | -0.0033 | -0.055 | 0.82 | 1.14 | 0.0362 | -0.037 | 4.58 |
| 114 | MAURITIUS | 5.40 | 4.49 | -0.0209 | -0.120 | 9.32 | 0.35 | -0.0032 | -0.143 | 5.83 |
| 115 | MAYOTTE | - | 5.46 | 0.0129 | -0.103 | - | 1.05 | 0.0065 | -0.154 | - |
| 116 | MEXICO | 2.03 | 0.86 | 0.1759 | -0.100 | 0.98 | 2.53 | 0.0117 | -0.109 | 5.37 |
| 117 | MOLDOVA | 2.03 | 1.03 | 0.0324 | -0.086 | 0.83 | 0.36 | -0.0037 | -0.127 | 6.60 |
| 118 | MONACO | 5.48 | 3.15 | -0.0044 | -0.147 | 1.66 | 0.54 | 0.0134 | -0.136 | 1.60 |
| 119 | MONGOLIA | 3.12 | 10.25 | 0.0116 | -0.204 | 1.98 | 0.68 | 0.0195 | -0.127 | 3.79 |
| 120 | MONTENEGRO | 8.16 | 1.37 | -0.0114 | -0.171 | 1.07 | 0.66 | 0.0040 | -0.085 | 8.42 |
| 121 | MOROCCO | 2.05 | 0.90 | 0.1161 | -0.114 | 0.84 | 0.95 | -0.0159 | -0.065 | 5.31 |
| 122 | MOZAMBIQUE | 2.14 | 0.72 | 0.0260 | -0.109 | 1.59 | 0.70 | 0.0152 | -0.068 | 8.17 |
| 123 | MYANMAR | 2.70 | 1.12 | -0.0028 | -0.113 | 0.83 | 1.15 | -0.0137 | -0.050 | 4.79 |
| 124 | NAMIBIA | 2.10 | 0.68 | 0.0315 | -0.049 | 1.03 | 1.22 | 0.0315 | -0.039 | 7.95 |
| 125 | NEPAL | 2.28 | 0.74 | 0.2070 | -0.035 | 0.91 | 0.78 | -0.0264 | -0.065 | 5.84 |
| 126 | NETHERLAND | 2.40 | 1.19 | 0.2485 | -0.043 | 0.92 | 1.04 | 0.0002 | -0.074 | 9.97 |
| 127 | NEW CALEDONIA | - | 5.00 | - | - | - | 1.00 | - | - | - |
| 128 | NEW ZEALAND | 5.63 | 0.74 | -0.0426 | -0.087 | 1.89 | 0.72 | 0.0140 | -0.099 | 9.21 |
| 129 | NICARAGUA | 5.76 | 0.97 | - | - | 1.39 | 1.02 | - | - | 8.56 |
| 130 | NIGER | 2.58 | 0.63 | -0.0231 | -0.083 | 0.96 | 2.21 | 0.0390 | -0.048 | 7.33 |
| 131 | NIGERIA | 1.91 | 1.13 | 0.0502 | -0.046 | 1.06 | 1.02 | 0.0333 | -0.047 | 3.89 |
| 132 | MACEDONIA | 1.84 | 0.74 | 0.0858 | -0.092 | 0.87 | 0.74 | 0.0528 | -0.230 | 6.58 |
| 133 | NORWAY | 2.40 | 0.77 | 0.2716 | -0.055 | 1.14 | 2.13 | 0.0052 | -0.145 | 10.05 |
| 134 | OMAN | 1.73 | 3.70 | 0.0972 | -0.092 | 1.13 | 9.80 | 0.0936 | -0.130 | 4.13 |
| 135 | PAKISTAN | 1.90 | 1.22 | 0.1301 | -0.060 | 1.02 | 1.19 | 0.0113 | -0.047 | 3.20 |
| 136 | PALESTINE | - | 0.96 | -0.0053 | -0.202 | - | 1.06 | 0.0063 | -0.050 | - |
| 137 | PANAMA | 2.08 | 0.96 | 0.1443 | -0.063 | 1.13 | 0.79 | 0.1195 | -0.070 | 7.27 |
| 138 | PAPAU NEW G. | 1.95 | 0.49 | -0.0081 | -0.115 | 2.45 | 0.88 | - | - | 2.37 |
| 139 | PARAGUAY | 2.22 | 0.59 | 0.0196 | -0.147 | 0.97 | 1.20 | 0.0032 | -0.168 | 6.65 |
| 140 | PERU | 2.35 | 0.89 | 0.0915 | -0.010 | 1.26 | 0.53 | -0.0077 | -0.111 | 5.24 |
| 141 | PHILLIPPINES | 2.29 | 1.15 | 0.1627 | -0.082 | 0.91 | 1.54 | 0.1772 | -0.174 | 4.40 |
| 142 | POLAND | 2.17 | 0.92 | 0.1562 | -0.079 | 0.99 | 1.31 | 0.0094 | -0.072 | 6.33 |
| 143 | POLYNESIA | - | 0.66 | - | - | - | 0.21 | - | -0.075 | - |
| 144 | PORTUGAL | 2.92 | 1.56 | 0.0301 | -0.140 | 1.15 | 3.89 | 0.0431 | -0.190 | 9.41 |
| 145 | QATAR | 2.61 | 0.80 | 0.0694 | -0.070 | 1.16 | 1.03 | -0.0019 | -0.094 | 2.49 |
| 146 | ROMANIA | 2.26 | 0.88 | 0.0218 | -0.056 | 0.91 | 0.95 | -0.0072 | -0.121 | 5.56 |
| 147 | RUSSIA | 2.41 | 1.07 | 0.0775 | -0.020 | 1.00 | 0.87 | 0.0046 | -0.037 | 5.32 |
| 148 | RWANDA | 2.03 | 1.80 | 0.0615 | -0.146 | 1.26 | 0.14 | 0.0382 | -0.064 | 7.54 |
| 149 | SAO TOME | 3.09 | 1.44 | -0.0218 | -0.153 | 3.33 | 2.67 | 0.0162 | -0.127 | 6.27 |
| 150 | SAN MARINO | 5.88 | 5.10 | -0.0157 | -0.137 | 1.14 | 0.26 | -0.0028 | -0.154 | 7.14 |
| 151 | SAUDI ARABIA | 2.31 | 0.90 | 0.0607 | -0.060 | 0.90 | 0.98 | -0.0138 | -0.029 | 6.36 |
| 152 | SENEGAL | 2.02 | 0.72 | 0.0351 | -0.047 | 1.24 | 1.59 | 0.0387 | -0.047 | 3.98 |
| 153 | SERBIA | 2.13 | 1.62 | 0.0042 | -0.053 | 0.79 | 0.82 | 0.0123 | -0.038 | 8.54 |
| 154 | SEYCHELLES | 2.68 | 0.48 | - | - | 1.94 | 0.54 | 0.0313 | -0.134 | 5.11 |
| 155 | SIERRA LEONE | 1.50 | 2.23 | 0.0143 | -0.107 | 1.52 | 1.37 | 0.0291 | -0.063 | 16.06 |
| 156 | SINGAPORE | 2.06 | 1.33 | 0.0551 | -0.030 | 1.52 | 2.83 | 0.0641 | -0.080 | 4.46 |
| 157 | SLOVAK | 1.74 | 0.99 | -0.0286 | -0.123 | 0.92 | 0.74 | 0.0028 | -0.193 | 6.69 |
| 158 | SLOVENIA | 1.78 | 0.75 | -0.0345 | -0.079 | 1.08 | 0.64 | -0.0004 | -0.263 | 8.30 |
| 159 | SOLOMON ISL. | - | - | - | - | - | 0.29 | - | - | 4.47 |
| 160 | SOMALIA | 1.95 | 1.18 | -0.0085 | -0.091 | 2.55 | 1.49 | - | - | - |
| 161 | SOUTH AFRICA | 2.54 | 0.87 | 0.257 | -0.110 | 1.15 | 1.72 | 0.0303 | -0.039 | 8.25 |
| 162 | SOUTH SUDAN | 2.99 | 0.58 | 0.0007 | -0.152 | 1.59 | 0.51 | 0.0095 | -0.133 | 6.40 |
| 163 | SPAIN | 3.85 | 0.38 | 0.3350 | -0.035 | 1.16 | 0.79 | 0.0029 | -0.080 | 8.98 |
| 164 | SRI LANKA | 4.14 | 2.13 | 0.0144 | -0.159 | 1.04 | 1.07 | 0.1347 | -0.160 | 3.76 |
| 165 | ST KITTS NEVIS | - | 2.00 | - | - | - | 1.00 | - | - | 5.31 |
| 166 | ST LUCIA | 1.34 | 1.13 | - | - | 2.86 | 0.69 | 0.0157 | -0.082 | 4.40 |
| 167 | ST VINCENT | 5.86 | 0.04 | - | - | 2.17 | 2.00 | 0.0407 | -0.080 | 4.47 |
| 168 | SUDAN | 1.97 | 0.36 | 0.0193 | -0.094 | 0.24 | 1.63 | 0.0407 | -0.039 | 4.51 |
| 169 | SURINAME | 1.41 | 10.34 | 0.0214 | -0.061 | 1.27 | 1.21 | 0.0379 | -0.042 | 7.97 |
| 170 | SWEDEN | 2.10 | 0.56 | 0.2572 | -0.106 | 1.05 | 0.28 | 0.0123 | -0.162 | 10.90 |
| 171 | SWITZERLAND | 2.86 | 1.21 | 0.2388 | -0.044 | 0.95 | 0.18 | -0.0082 | -0.041 | 11.88 |
| 172 | SYRIA | 2.80 | 1.43 | 0.0311 | -0.030 | 1.06 | 0.66 | 0.0086 | -0.041 | - |
| 173 | TAIWAN | 3.42 | 1.88 | 0.0036 | -0.084 | 1.84 | 1.49 | 0.0053 | -0.123 | - |
| 174 | TAJIKISTAN | 1.68 | 1.02 | 0.0418 | -0.066 | 0.54 | 1.89 | -0.0016 | -0.131 | 7.24 |
| 175 | TANZANIA | 5.00 | 0.91 | 0.1205 | -0.125 | 18.4 | - | - | - | 3.63 |
| 176 | THAILAND | 3.42 | 0.69 | -0.0201 | -0.055 | 1.63 | 2.71 | 0.0496 | -0.100 | 3.79 |
| 177 | TIMOR LESTE | - | 5.00 | - | - | - | 1.33 | - | - | 4.33 |
| 178 | TOGO | 2.09 | 0.08 | 0.0093 | -0.084 | 1.41 | 1.14 | 0.0083 | -0.112 | 6.17 |
| 179 | TRINIDAD | 5.35 | 0.32 | -0.0025 | -0.139 | 1.30 | 0.55 | -0.0102 | -0.113 | 6.93 |
| 180 | TUNISIA | 2.64 | 1.53 | -0.0122 | -0.084 | 1.15 | 2.77 | 0.0053 | -0.117 | 7.29 |
| 181 | TURKEY | 4.32 | 1.15 | 0.0120 | -0.040 | 0.81 | 2.21 | 0.0078 | -0.030 | 4.12 |
| 182 | UAE | 2.33 | 0.97 | 0.0484 | -0.080 | 1.22 | 1.15 | 0.0085 | -0.055 | 4.23 |
| 183 | UGANDA | 2.18 | 0.95 | - | - | 0.88 | 0.64 | 0.0047 | -0.154 | 6.53 |
| 184 | UKRAINE | 2.16 | 0.96 | 0.0325 | -0.130 | 0.89 | 0.30 | -0.0032 | -0.093 | 7.72 |
| 185 | UK | 2.89 | 0.76 | 0.2223 | -0.037 | 1.25 | 1.03 | 0.0106 | -0.035 | 10.00 |
| 186 | USA | 3.85 | 8.42 | 0.2882 | -0.030 | 0.99 | 0.49 | 0.0121 | -0.060 | 16.89 |
| 187 | URUGUAY | 2.76 | 0.63 | -0.0228 | -0.086 | 1.15 | 1.03 | 0.0389 | -0.039 | 9.20 |
| 188 | UZBEKISTAN | 1.82 | 0.95 | 0.1231 | -0.088 | 0.71 | 0.90 | 0.0238 | -0.170 | 5.29 |
| 189 | VENEZUELA | 2.57 | 1.54 | 0.0389 | -0.073 | 0.94 | 0.82 | 0.0002 | -0.134 | 3.56 |
| 190 | VIETNAM | 3.59 | 3.29 | -0.0166 | -0.158 | 1.94 | 1.43 | -0.0040 | -0.158 | 5.92 |
| 191 | VIRGIN ISLANDS | - | 0.51 | - | - | - | 0.33 | - | - | - |
| 192 | WEST GAZA | 3.73 | 1.00 | - | - | 0.87 | 0.98 | - | - | - |
| 193 | YEMEN | 1.57 | 0.70 | 0.0049 | -0.164 | 2.84 | 1.50 | 0.0006 | -0.150 | - |
| 194 | ZAMBIA | 2.80 | 0.75 | 0.0265 | -0.134 | 1.73 | 1.12 | 0.0372 | -0.046 | 4.93 |
| 195 | ZIMBABWE | 1.98 | 1.44 | 0.0367 | -0.087 | 1.40 | 1.62 | 0.0438 | -0.045 | 4.73 |

## Appendix B

### Abbreviations

ARIMAF: Opposite autocorrelation slope for first wave
ARIMAS: Opposite autocorrelation slope for second wave
SecondR: Maximum R_0_ for second wave from [3]
SecondD: Deterministic R_0_ for second wave from [2]
FirstwaveR: Maximum R_0_ for first wave from [3]
FirstwaveD: Deterministic R_0_ for second wave from [2]
CHE/GDP: Current Health Expenditure as Gross Domestic Product percentage [1]

## Author Contributions

All authors contributed equally in the development and read the final version of the paper.

## Acknowledgments

The authors wish to acknowledge the Petroleum Technology Development Fund (PTDF) Nigeria doctoral fellowship in collaboration with Campus France Africa Unit.

## References

1. Worldbank. Available online: https://data.worldbank.org/indicator/SH.XPD.CHEX.GD.ZS (accessed on line on 12th February 2021) 2021.

2. Demongeot, J.; Oshinubi, K.; Seligmann, H.; Thuderoz, F. Estimation of Daily Reproduction Rates in COVID-19 Outbreak. MedRxiv 2021, doi.org/10.1101/2020.12.30.20249010.

3. Worldometers. Available online: https://www.worldometers.info/coronavirus/ (accessed on 12th February 2021) 2021.

4. Renkulab. Available online: https://renkulab.shinyapps.io/COVID-19-Epidemic-Forecasting/_w_e213563a/?tab=ecdc_pred&country=France (accessed on 22nd February 2021) 2021.

5. Wiener, N. Extrapolation, Interpolation, and Smoothing of Stationary Time Series; The MIT Press: Cambridge (Mass.) 1949.

6. Pedregosa, F.; Varoquaux, G.; Gramfort, A.; Michel, V.; Thirion, B.; Grisel, O.; Blondel, M.; Prettenhofer, P.; Weiss, R.; Dubourg, V.; Vanderplas, J.; Passos, A.; Cournapeau, D.; Brucher, M.; Perrot, M. E. Duchesnay, E.. Scikit-learn: Machine Learning in Python. Journal of Machine Learning Research 2011, 12, 2825–2830.

7. Demongeot, J.; Flet-Berliac, Y.; Seligmann, H. Temperature decreases spread parameters of the new covid-19 cases dynamics. Biology (Basel) 2020, 9, 94.

8. Seligmann, H.; Iggui, S.; Rachdi, M.; Vuillerme, N.; Demongeot, J. Inverted covariate effects for mutated 2nd vs 1st wave Covid-19: high temperature spread biased for young. Biology (Basel) 2020, 9, 226.

9. Demongeot, J.; Griette, Q.; Magal, P. Computations of the transmission rates in SI epidemic model applied to COVID-19 data in mainland China. Royal Society Open Science 2020, 7, 201878.

10. Soubeyrand, S.; Demongeot, J.; Roques, L. Towards unified and real-time analyses of outbreaks at country-level during pandemics. One Health 2020, 11, 100187.

11. Seligmann, H.; Vuillerme, N.; Demongeot, J. Unpredictable, Counter-Intuitive Geoclimatic and Demographic Correlations of COVID-19 Spread Rates. Biology (Basel) 2021, 10, 623.

12. Gaudart, J.; Landier, J.; Huiart, L.; Legendre, E.; Lehot, L.; Bendiane, M.K.; Chiche, L.; PetitJean, A.; Mosnier, E.; Kirakoya-Samadoulougou, F.; Demongeot, J.; Piarroux, R.; Rebaudet, S. Factors associated with spatial heterogeneity of Covid-19 in France: a nationwide ecological study. The Lancet Public Health 2021, 6, e222–e231.

13. Griette, Q.; Demongeot, J.; Magal, P. A robust phenomenological approach to investigate COVID-19 data for France. Math. Appl. Sci. Eng. 2021, 2, doi.org/10.5206/mase/14031.

